# Fasting Improves Treatment Efficacy in Cancer Patients Without Compromising Safety

**DOI:** 10.1101/2025.04.18.25326094

**Authors:** Zeyao Wang, Xueying Wang, Lina Cui, Zihan Liu, Fanxuan Huang, Zhaoyu Pan, Jiaqing Xiao, Tong Liu

## Abstract

The vulnerability of cancer cells to nutrient deprivation and their reliance on specific metabolites are emerging markers in cancer research. Numerous animal studies and preclinical research have shown that fasting is both safe and feasible. As a result, many studies suggest combining fasting with chemotherapy, immunotherapy, or other treatments as a potentially promising strategy to enhance therapeutic outcomes. This article provides a comprehensive analysis of the outcome measures in randomized controlled trials that reflect the impact of fasting on cancer treatment efficacy. Additionally, it validates the relationship between fasting and relevant proteins using data from the UK Biobank, including insulin receptor (INSR), insulin-like growth factor 1 receptor (IGF1R), and insulin-like growth factor binding protein 1 (IGFBP1), and further examines the impact of changes in these proteins on cancer patient prognosis. We found that fasting improves radiological response rates by lowering glucose, insulin, and insulin-like growth factor-1 (IGF-1) levels, while increasing insulin-like growth factor binding protein (IGF-BP) levels. Fasting also reduces DNA damage in immune cells and does not lead to additional treatment-related toxicities. Thus, fasting is a safe and effective adjunctive treatment for cancer, improving therapeutic outcomes while ensuring patient safety.

## 1. Introduction

Cancer remains a formidable public health challenge, consistently ranking as the second leading cause of morbidity and mortality in the United States(1). Recent projections indicate that more than 2 million individuals will face a cancer diagnosis in 2024, with over 600,000 likely to succumb to the disease. (https://seer.cancer.gov/statfacts/html/all.html). With advancements in cancer treatment, chemotherapy, radiotherapy, and immunotherapy have become standard approaches. However, these treatments often cause side effects due to damage to normal tissues(2). Moreover, current treatments often fail to achieve ideal prognoses for patients (3, 4). Recent studies suggest that environmental factors, particularly dietary factors, may play a more significant role in cancer development(5, 6). As a result, fasting, which controls dietary intake, has been hypothesized to prevent cancer and improve the efficacy and tolerability of anticancer therapies(7–9). Preclinical models have shown that fasting can reduce damage to normal cells, inhibit cancer cell proliferation and metastasis, and increase cancer cell death (10–13). This is because fasting alters cellular metabolic pathways, placing cells in a nutrient-deprived environment. In normal cells, fasting reduces nutrient intake, which lowers glucose levels, inhibits the PKA-AMPK-EGR1 pathway (14–16), enabling normal cells to adapt to fasting-induced metabolic stress, regulate energy metabolism-related genes, and enhance antioxidant genes (e.g., SOD, CAT) and anti-apoptotic genes (e.g., Bcl-2) to protect against oxidative stress damage. Additionally, fasting decreases IGF-1 levels, inhibiting the PI3K-Akt-mTOR signaling pathway and promoting the expression of protective and antioxidant genes (17–19). These mechanisms protect normal cells, reduce treatment-related side effects, and improve patient outcomes and survival (12, 20–22). In contrast, cancer cells are unable to effectively adapt to nutrient deprivation, increasing their sensitivity to therapy (8, 9, 23). However, clinical models investigating fasting as an adjunct to cancer treatment remain limited. To further investigate the impact of fasting on cancer treatment efficacy, this article performs a meta-analysis of randomized controlled trials (RCTs) focusing on key outcome measures including radiological response, immune cell DNA damage, changes in metabolic parameters, treatment-related toxicity, and quality of life scores. We also analyze data from the UK Biobank (UKB) Olink database, examining the relationship between fasting duration and the levels of metabolic and hematological proteins, such as insulin receptor (INSR), insulin-like growth factor 1 receptor (IGF1R), insulin-like growth factor binding protein 1 (IGFBP1), transferrin (TF), hemoglobin subunits (HBQ1, HBZ), platelet glycoproteins (GP1BA, GP1BB), granulocyte colony-stimulating factor (CSF3), interleukin-8 (CXCL8), myeloperoxidase (MPO), and tumor necrosis factor-alpha (TNF), and how these factors relate to cancer patient prognosis. In conclusion, fasting can significantly improve cancer treatment efficacy without increasing the incidence of treatment-related adverse effects or hematologic toxicity. In the future, fasting may be integrated with existing cancer therapies, becoming a safe and effective adjunctive treatment.

## 2. Materials and Methods

This article summarizes the results of multiple relevant RCTs and integrates Olink data from the UK Biobank to explore the impact of fasting on the effectiveness of cancer treatment.

### 2.1. Meta-analysis

According to the Preferred Reporting Items for Systematic Reviews and Meta-Analyses (PRISMA) guidelines, a systematic approach was used to retrieve and evaluate the literature(24). Electronic databases such as PubMed (MEDLINE), Web of Science, Embase, and Cochrane Library (CENTRAL) were used to select studies included in this systematic review and meta-analysis. The following search terms were employed: “fasting,” “cancer,” and “RCT,” using a combination of subject headings and free-text keywords (Table S1). The literature search was completed on November 20, 2024. The protocol was prospectively registered in the International Prospective Register of Systematic Reviews (PROSPERO) (CRD42024617851) (https://www.crd.york.ac.uk/PROSPERO/) (accessed on December 5, 2024). A total of 4877 unique records were identified through the database and bibliographic searches. Of these, 4869 were excluded based on the title, abstract content, and full texts by two researchers,with any discrepancies discussed until a consensus was reached; thus, only eight records met inclusion criteria (a flow diagram of the review process is shown in Supplementary Figure 12).

#### 2.1.1 Inclusion and Exclusion Criteria

The inclusion and exclusion criteria for the article were established based on the PICOS principle:(1)Population: Patients who meet the diagnostic criteria for cancer;(2) Intervention/exposures: Fasting including short-term fasting, fast-mimicking diet, intermittent fasting and short-term caloric restriction diet;(3) Comparison: A placebo-controlled intervention or no intervention;(4) Outcomes: Radiological response, DNA damage levels, metabolic parameters, adverse events, hematological parameters, and quality of life scores;(5) Type of study: Randomized controlled trials. Articles were excluded if they were not receiving standard care for their malignancies; had no comparison group; had no exposure or outcome of interest; were non-randomized; were non-human species; were conference abstracts, book chapters, reviews, or other forms without detailed empirical data.

#### 2.1.2 Data extraction

Based on the research objectives and outcome measures, the following information was independently collected by two reviewers from published clinical trials and data in the literature (Table 1): (1) first author and publication year; (2) study design; (3) cancer type; (4) intervention; (5) intervention duration; (6) treatment therapy; (7) comparison; (8) outcome.

**Table 1:**
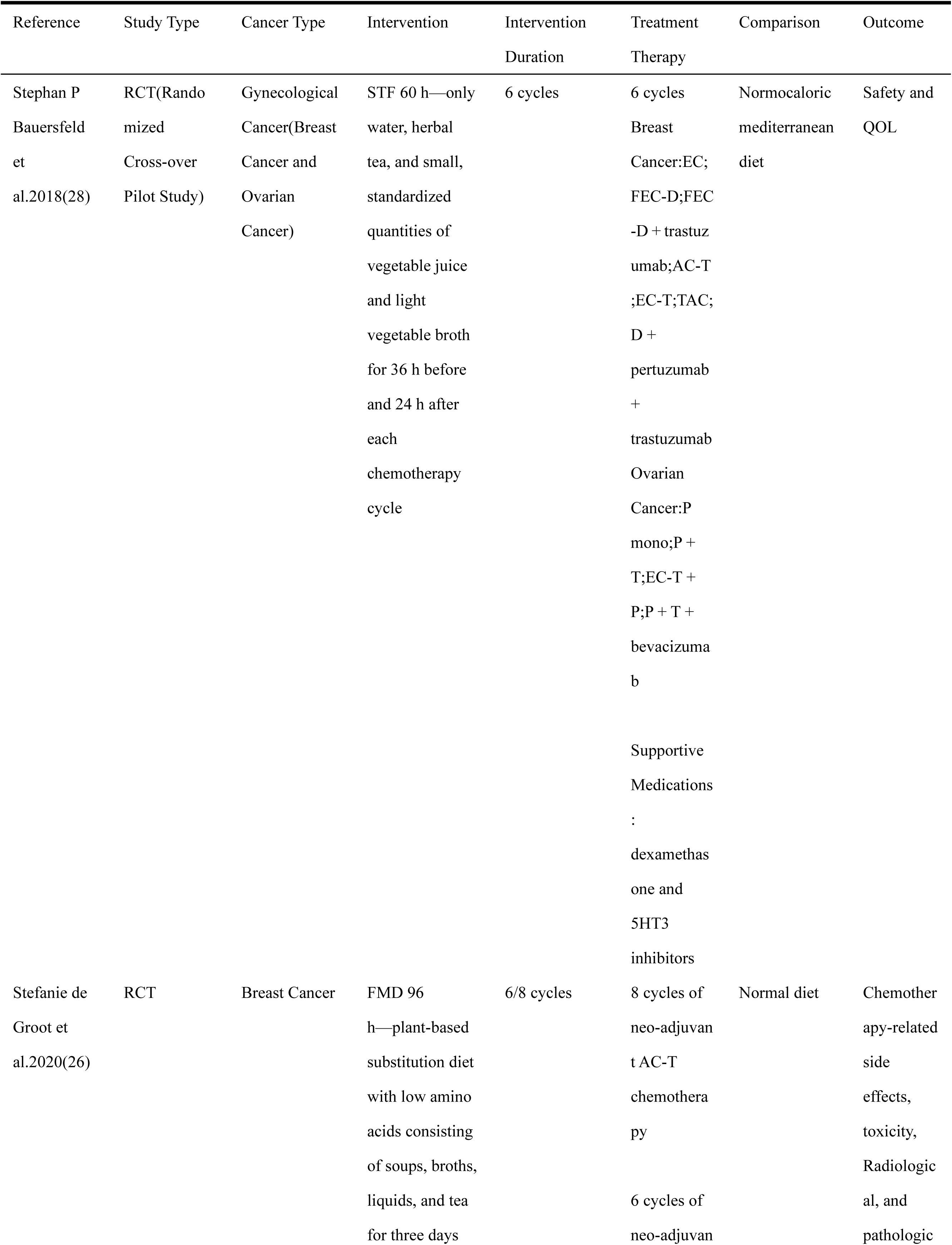
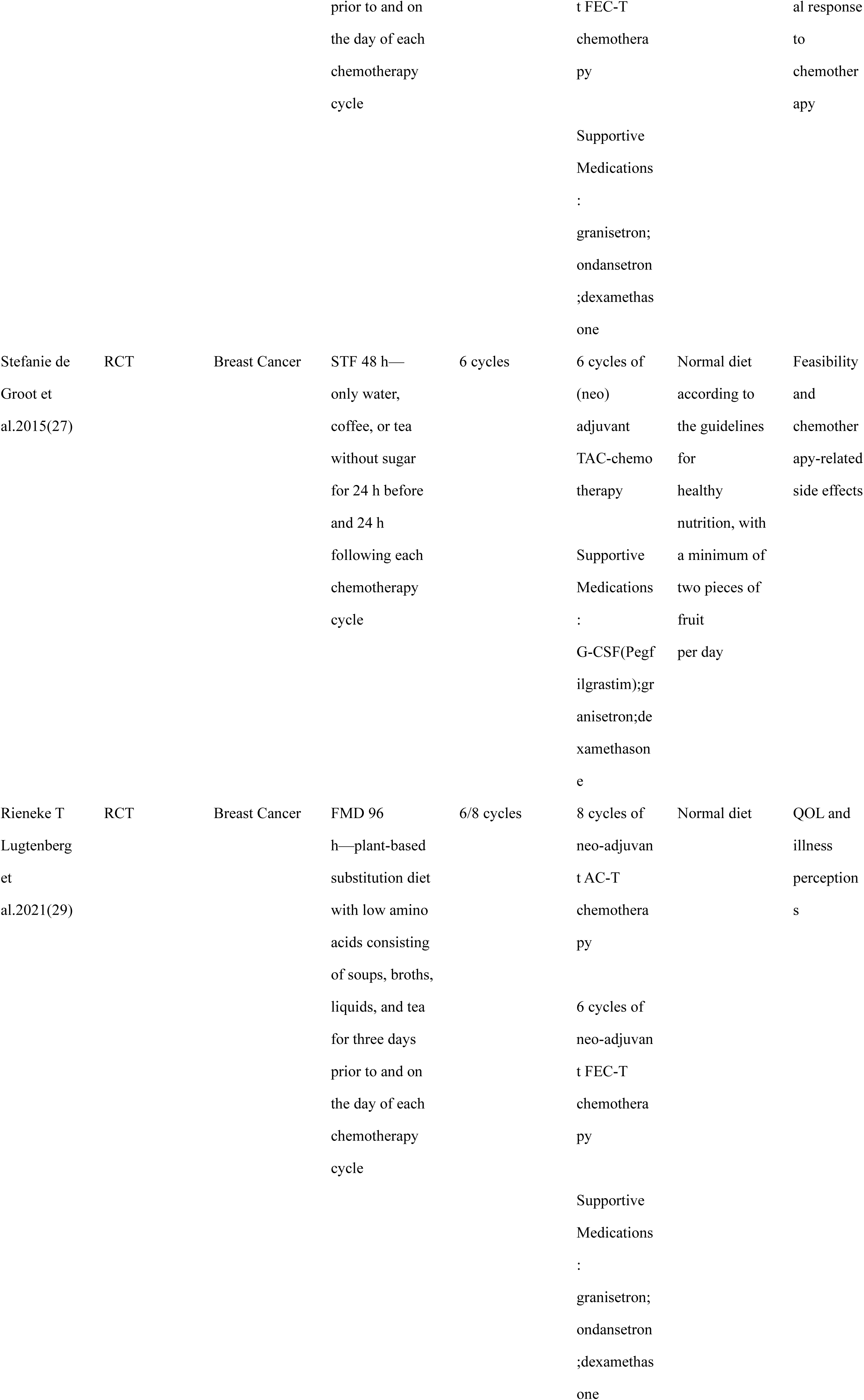
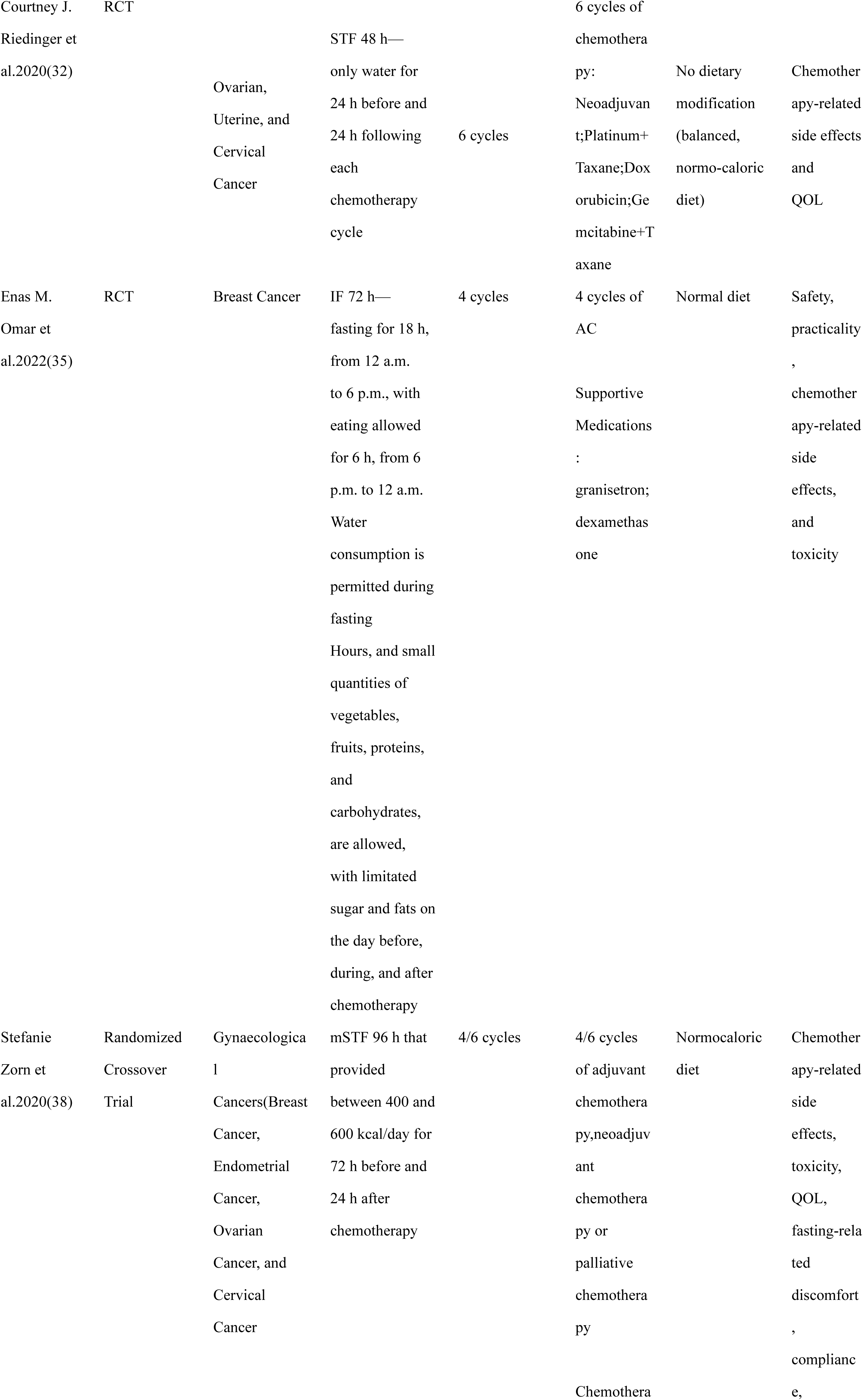
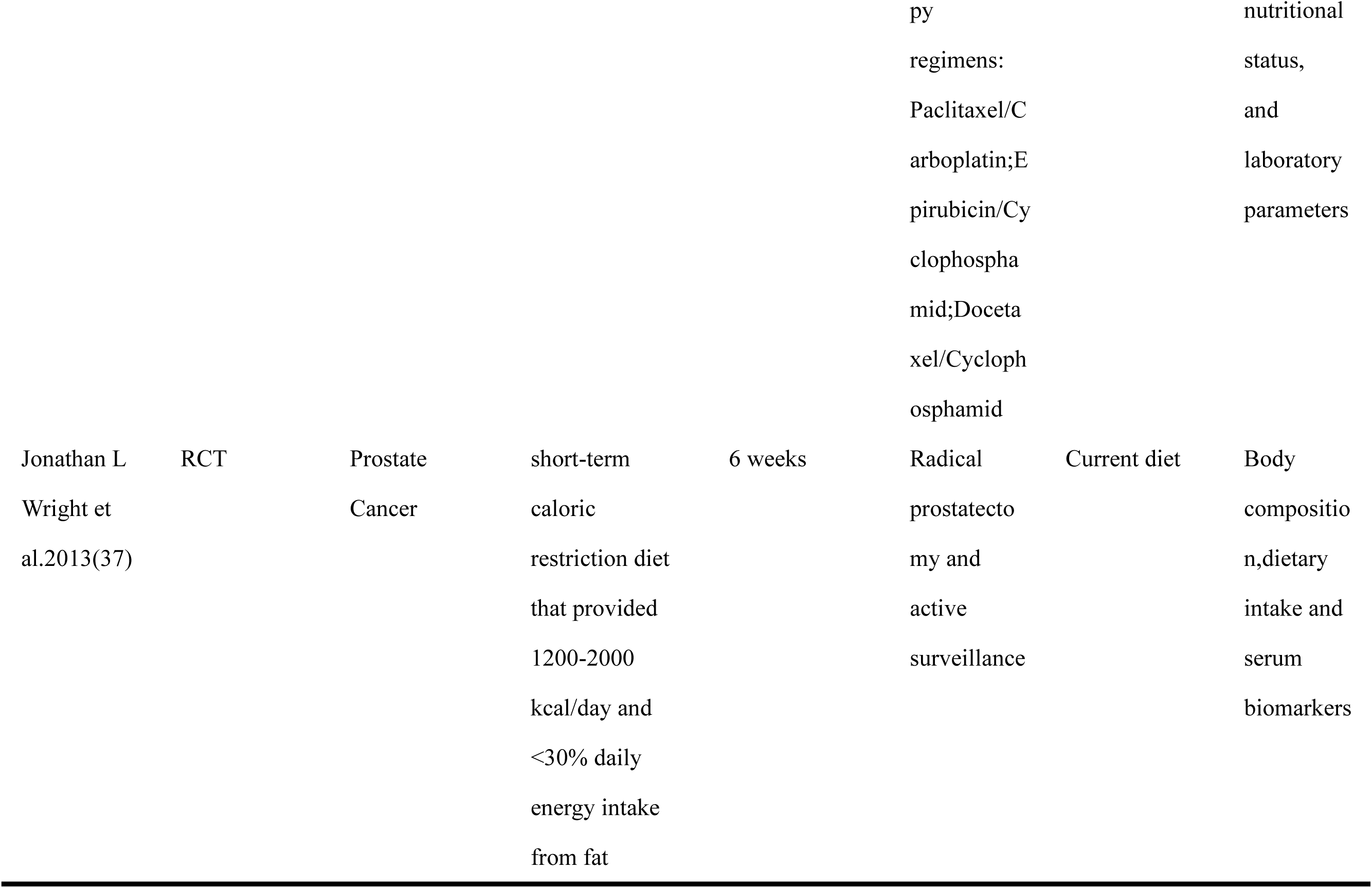
Basic characteristics of the included studies.

#### 2.1.3 Outcome

The results of this article are mainly divided into the following five parts: (1) Radiological response: Radiological response, according to the Miller and Payne classification, is divided into four categories: Complete Response (CR), Partial Response (PR), Stable Disease (SD), and Progressive Disease (PD). We used the RECIST 1.1 criteria(25) to assess clinical response through imaging at both the midpoint and end of treatment, with CR and PR being used to represent effective tumor treatment. (2) γ-H2AX intensity: According to Stefanie de Groot et al.2020 (26) and Stefanie de Groot et al.2015 (27) ,PBMCs were isolated using Ficoll-Amidotrizoaat gradient centrifugation according to the standard operating procedure of the Medical Oncology department of LUMC. Isolated PBMCs were carefully resuspended and 3 times washed in PBS. Samples were fixed in 1.5% formaldehyde and permealized in ice-cold pure methanol. Cells were washed 3 times in staining buffer (PBS with 5% bovine serum albumin) and stained for 30 min on ice with anti-CD45-PerCP-Cy5.5, clone 2D1 anti-CD3-PE (BD, clone SK7), anti-CD14-AF700 (BD, clone M5E2), anti-CD15-PE CF594 (BD, clone W6D3) and anti-γ-H2AX-AF488(Biolegend, clone 2F3), followed by another washing step and resuspension in PBS. The cell acquisition was performed immediately after the staining procedure on the flow cytometer and data were analyzed using BD FACS Diva Software version 6.2. The CD45+ cells were gated, after which the CD3+ T-lymphocytes or CD14+ CD15− monocytes were analyzed for the geomean (as measure for the intensity) of γ-H2AX. (3) Metabolic and hematological parameters levels: After treatment, venous blood was drawn from patients to measure the levels of metabolic parameters, such as glucose, insulin, IGF-1, and IGF-BP, along with hematological parameters. (4) Adverse events: The adverse events reported in each study were graded according to the Common Terminology Criteria for Adverse Events (CTCAE). The primary outcomes were grade III/IV total adverse events, grade III/IV neutropenia, and grade III/IV neutropenic fever. (5) Scale Scores: For measuring health-related QOL we used the functional assessment of chronic illness therapy (FACIT©) measurement system according to Stephan P Bauersfeld et al.2018 (28). The Functional Assessment of Cancer Therapy-General (FACT-G©) forms the generic core questionnaire of all FACIT© scales. The FACIT© scales are constructed to complement the FACT-G© scale by addressing relevant disease-, treatment-, or condition-related issues not already covered in the general questionnaire. The Trial Outcome Index (TOI) is a measure of physical aspects of QOL. It is the sum of the FACT-G subscales of physical well-being (PWB), functional well-being (FWB), and any FACIT© disease-, treatment-, or condition-specific scale. For additional concerns we used the FACIT-F©, a 13-item questionnaire that assesses self-reported fatigue and its impact upon daily activities. Altogether we obtained 8 different scales and subscale scores: the subscales PWB, EWB, SWB, FWB as compounds of the FACT-G© scale (27 items); the additional subscale FACIT-F© (13 items); the TOI-FACIT-F© (27 items) and the total FACIT-F© scale as union of the FACT-G© and FACIT-F© scales (40 items). In addition, according to Stefanie de Groot et al.2020(26) and Rieneke T Lugtenberg et al.2021(29), we assessed global health status using the EORTC QLQ-C30 (30) before treatment (post-randomization), during treatment, at the end of treatment, and at the 6-month follow-up. Higher functional scale scores (ranging from 0 to 100) indicate better quality of life (QoL). Psychosocial distress was measured using a scale ranging from 0 (no distress) to 10 (extreme distress). Patients were asked to circle the number that best describes the overall distress they experienced in the past week at mid-treatment, end of treatment, and the 6-month follow-up, according to Stefanie de Groot et al.2020(26) and Rieneke T Lugtenberg et al.2021(29).

#### 2.1.4 Quality assessment

The risk of bias for the included studies was evaluated using the Cochrane Handbook for Systematic Reviews of Interventions’ recommended bias risk assessment tool. This tool includes 7 key items: random sequence generation, allocation concealment, blinding of participants and interventions, blinding of outcome assessors, completeness of outcome data, reporting bias, and other biases. The assessment results were classified as high risk, low risk, or unclear risk. Risk of bias assessment plots were generated using RevMan 5.4 software (Supplementary Figure 10 and 11). Additionally, the Critical Appraisal Skills Programme (CASP) was used to assess the quality of the randomized controlled trials (Table S2). Literature search, screening, data extraction, and quality assessment were independently performed by two researchers, with the results being compared and discussed. In case of any discrepancies, a third researcher was consulted to finalize the results.

### 2.2 UK Biobank database

We used Olink data from the UK Biobank to analyze the relationship between fasting duration in cancer patients and the expression levels of metabolic and hematological parameters, as well as the association between these protein expression levels and overall survival (OS). A total of 1,862 participants from the UK Biobank were included in this study. These patients were diagnosed with one or more cancers between 2006 and 2010, with documented fasting durations. First, to analyze the correlation between fasting duration and protein expression levels, we used Spearman’s rank correlation analysis, as the data for fasting duration and protein expression levels were ordered and did not meet the normal distribution assumption. The Spearman test is suitable for evaluating monotonic relationships, especially for data with skewed distributions or nonlinear associations. To further determine whether there were differences in protein expression levels between fasting and satiety groups, we defined the fasting group as those with a fasting duration of ≥12 hours, and the satiety group as those with a fasting duration of ≤6 hours. We excluded patients with fasting durations between 6 and 12 hours from the differential analysis, resulting in 1,798 patients being included. The comparison of protein expression levels between the fasting and satiety groups was performed using the Mann-Whitney U test, as the protein expression data did not meet the normal distribution assumption and consisted of independent samples. Nonparametric rank-sum tests do not require the assumption of normality, making them suitable for skewed data distributions. Finally, we obtained the OS for the 1,862 patients initially included in the UK Biobank based on their diagnosis and death dates. Since a single patient may have been diagnosed with different cancers at different times between 2006 and 2010, 2,642 data points were included in the survival analysis. For patients in the UK Biobank with no recorded death dates, we used the follow-up end time and recorded the survival status for each patient (1 for death, 0 for survival). Protein expression levels were divided into high and low expression groups based on the data distribution, and the Kaplan-Meier method was used to generate survival curves to visually compare the survival differences between the high and low protein expression groups.

### 2.3 Statistical analysis

Data analysis was conducted using RevMan 5.4 and STATA 18 software. For dichotomous data, the relative risk (RR) was used as the effect size, while for continuous variables, the mean difference (MD) was calculated, along with 95% confidence intervals (CIs) and p-values. Heterogeneity was assessed using both P-values and the *I²* statistic. When *I²* ≤ 50% and P ≥ 0.10, the heterogeneity was considered low, and a fixed-effects model was applied. When *I²* > 50% and/or P < 0.10, significant heterogeneity was considered, and sensitivity analysis was performed to explore the sources of heterogeneity. If heterogeneity could not be reduced, a random-effects model was used. Funnel plots were created to assess publication bias for outcomes with at least five included studies, while Begg’s and Egger’s tests were conducted for outcomes with at least three included studies.

## 3. Results

### 3.1 Fasting enhances the radiographic response rate in cancer patients

Radiographic response is an important indicator for evaluating the efficacy of cancer treatments, such as chemotherapy, targeted therapy, and immunotherapy, by assessing tumor shrinkage or stability (31). To explore the impact of fasting on the rates of complete response (CR) and partial response (PR), we included two studies (26, 32). Heterogeneity testing showed no significant statistical heterogeneity between the two studies (*I²*=0, P=0.93) (Figure 1). Therefore, a fixed-effects model was used for analysis. The results revealed that the CR and PR rates in the fasting group were significantly higher than those in the normal diet group (RR=1.22, 95% CI [1.03, 1.44], P=0.02). This suggests that fasting may enhance the therapeutic response and improve treatment outcomes, providing new evidence for its potential role as an adjunct in cancer therapy.

**Figure 1:**
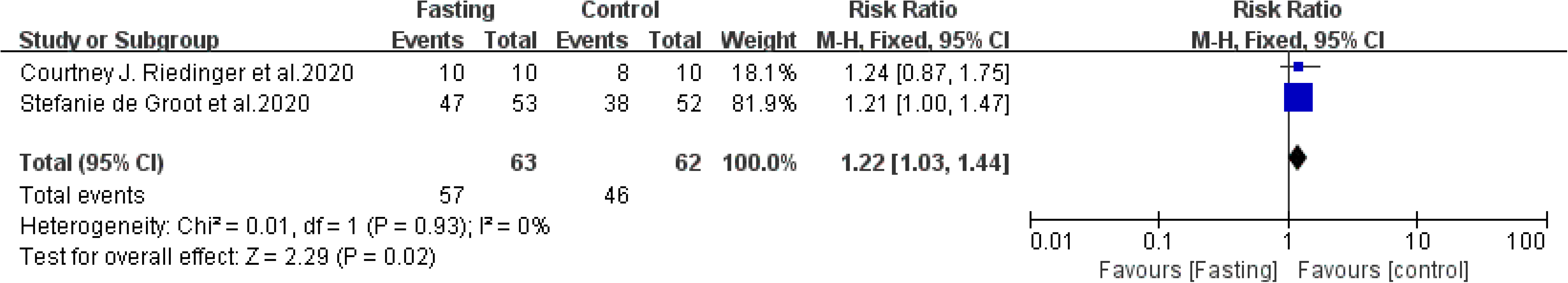
Forest plot in radiological response Stefanie de Groot et al.2020, Courtney J. Riedinger et al.2020 CI, confidence interval; *I2* measure of heterogeneity

### 3.2 Fasting reduces DNA damage in immune cells of cancer patients

Phosphorylation of γ-H2AX is a well-established marker for DNA double-strand breaks and is widely used to assess the extent of DNA damage in cells (33). In cancer treatment, the level of DNA damage in immune cells directly influences the therapeutic efficacy. Therefore, studying the γ-H2AX intensity in immune cells can provide insights into the potential impact of fasting on cancer treatment outcomes. This study included two studies(26, 27), which compared the γ-H2AX expression levels in CD45+CD3+ T lymphocytes and CD45+CD14+CD15-monocytes between the experimental and control groups. Heterogeneity testing showed no significant statistical heterogeneity between the studies (*I²*=24%, P=0.25; *I²*=0, P=0.6) (Figure 2), thus a fixed-effects model was applied for further analysis. The results indicated that there was no significant difference in the γ-H2AX intensity in CD45+CD3+ T lymphocytes between the fasting and normal diet groups (MD=-9.95, 95% CI [-25.92, 6.03], P=0.22). However, the γ-H2AX intensity in CD45+CD14+CD15-monocytes was significantly lower in the fasting group compared to the normal diet group (MD=-53.56, 95% CI [-89.59, -17.53], P=0.004). This finding suggests that fasting may reduce DNA damage in immune cells, particularly in monocytes during treatment, thereby indirectly enhancing anti-tumor immune responses and improving the overall efficacy of cancer therapy.

**Figure 2:**
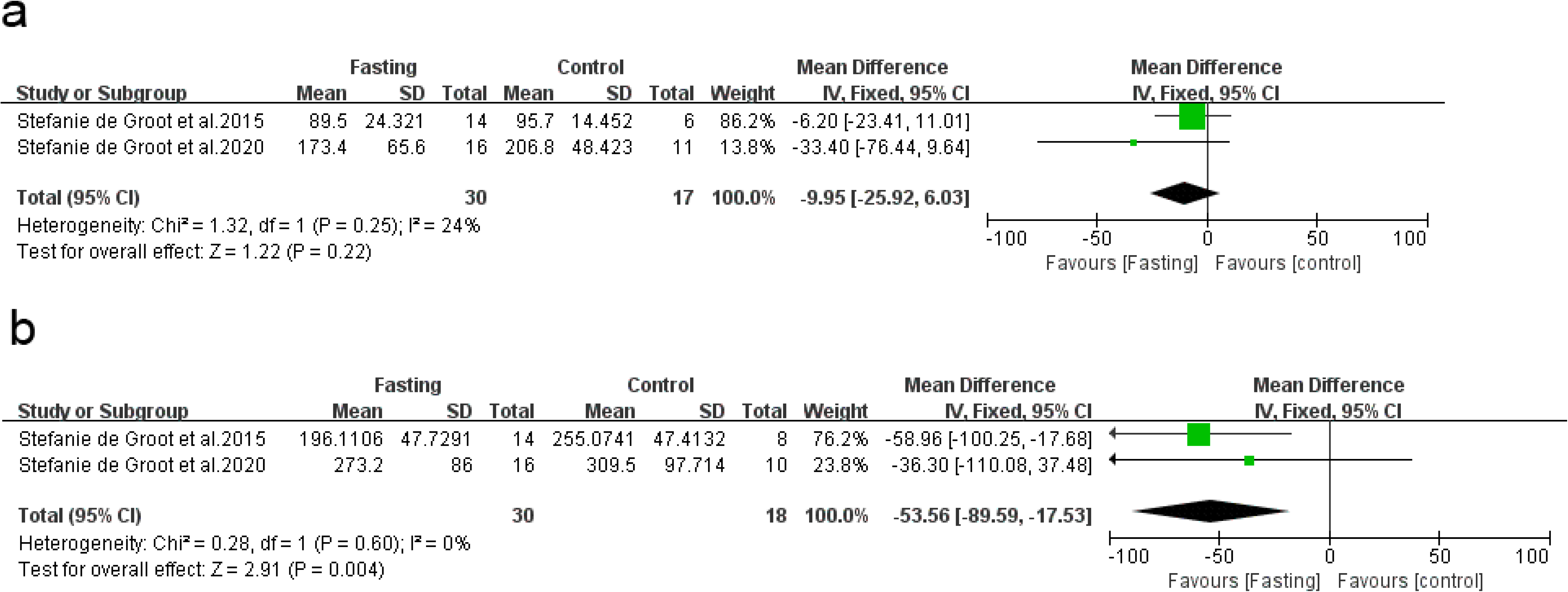
Forest plot in γ-H2AX intensity Stefanie de Groot et al.2020, Stefanie de Groot et al.2015 CI, confidence interval; *I*^2^ measure of heterogeneity

### 3.3 Fasting enhances metabolic activity

Glucose levels are a key indicator of metabolic activity, particularly in cancer patients, where alterations in glucose metabolism may be closely associated with cancer onset, progression, and treatment response (34). Three studies (27, 32, 35) included in our analysis reported changes in glucose levels between the fasting and normal diet groups. Initial heterogeneity tests revealed significant variability across the studies (*I²*=65%, P=0.06) (Supplementary Figure 1a). To further investigate the source of heterogeneity, we performed sensitivity analysis by excluding each study one by one. The results indicated that the study by Enas M. Omar et al. 2022(35) had a significant impact on the stability of the results (Supplementary Figure 2a). A deeper analysis (Table 1) revealed that this instability was likely due to differences in the fasting protocols, specifically variations in daily calorie restriction standards. Based on this finding, we excluded this study and performed a reanalysis, which showed that the heterogeneity was no longer statistically significant (*I²*=18%, P=0.27) (Figure 3a).Using a fixed-effect model for the data analysis, we found that glucose levels in the fasting group were significantly lower than those in the normal diet group (MD=-16.67, 95% CI [-32.84, -0.67], P=0.04). This suggests that fasting may significantly reduce glucose levels by modulating metabolic pathways, potentially exerting a beneficial effect on tumor growth and treatment outcomes.

**Figure 3:**
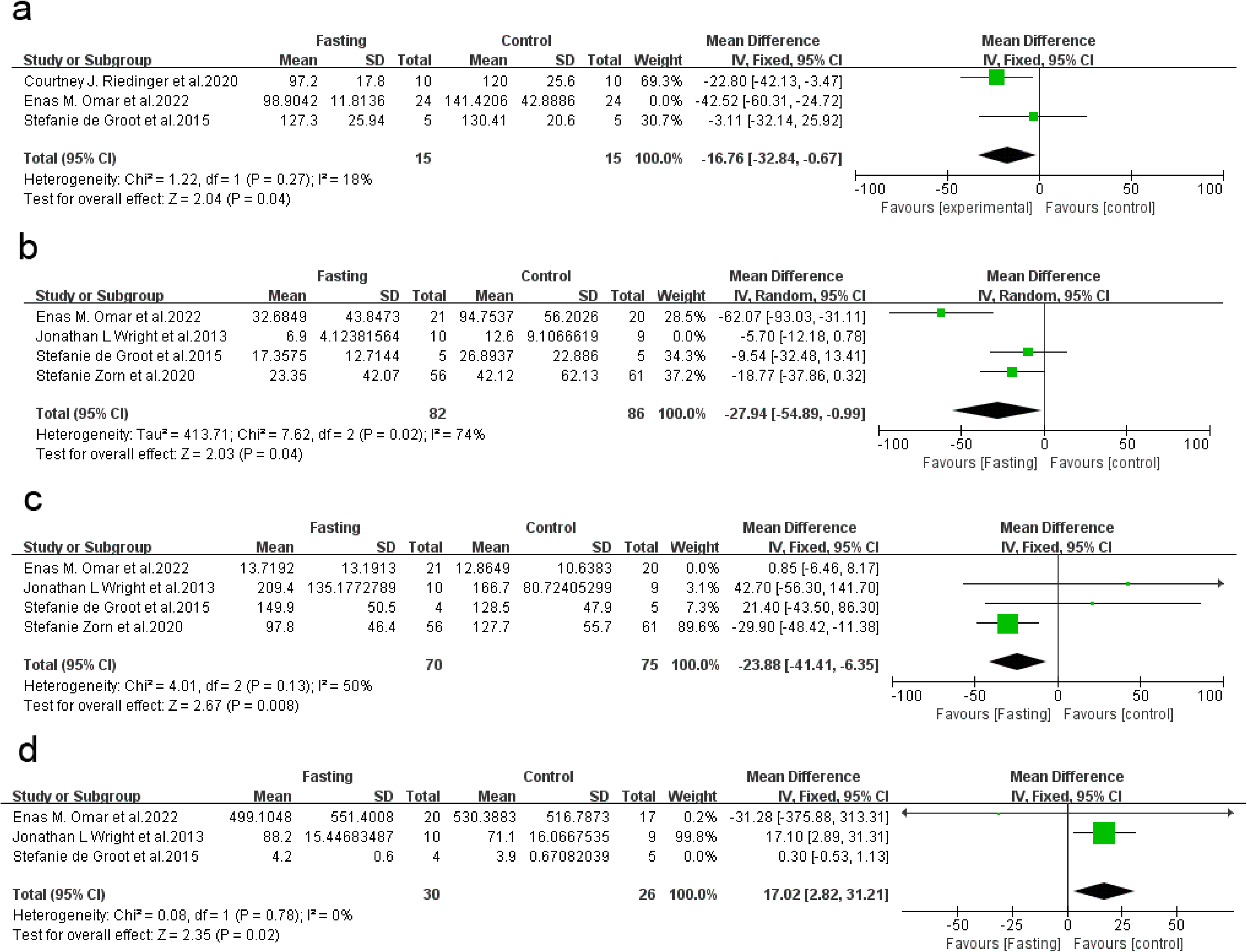
Forest plot in metabolic parameters(after) Courtney J. Riedinger et al.2020, Enas M. Omar et al.2022, Stefanie de Groot et al.2015, Stefanie Zorn et al.2020, Jonathan L Wright et al.2013 CI, confidence interval; *I*^2^ measure of heterogeneity

Insulin plays a crucial role in maintaining energy balance in the body, particularly in the metabolism of cancer patients(36). Changes in insulin levels are not only important indicators of metabolic health but may also be closely associated with the efficacy of cancer treatments. Four studies (27, 35, 37, 38) have analyzed the effect of fasting interventions on insulin levels. Initial heterogeneity tests revealed significant variability across three studies (*I²*=77%, P=0.004) (Supplementary Figure 1b). To further investigate the source of this heterogeneity, we conducted a sensitivity analysis and found that the study by Jonathan L Wright et al. 2013 (37) had a substantial impact on the stability of the results (Supplementary Figure 2b). Based on the characteristics of this study (Table 1), we reasonably hypothesize that two factors contributed to the instability of the results: first, the study enrolled a specific population of overweight or obese prostate cancer patients; second, the daily caloric intake in the fasting intervention differed—this study required 1200-2000 kcal per day for the experimental group, whereas the other three studies had much lower caloric intake thresholds. After excluding this study, we conducted another heterogeneity test, but the result still showed significant variability (*I²*=74%, P=0.02) (Figure 3b). Further analysis indicated that this heterogeneity was likely due to differences in fasting protocols between the study by Enas M. Omar et al. 2022 and the other studies. Therefore, the heterogeneity of the data is unavoidable and is a common phenomenon in studies with different designs. To mitigate this, we applied a random effects model for data analysis, which showed that insulin levels were significantly lower in the fasting group compared to the normal diet group (MD=-27.94, 95%CI [-54.89, -0.99], P=0.04). This result suggests that fasting can significantly modulate insulin levels, providing new evidence for its potential as an adjunctive treatment in cancer therapy. To validate the clinical significance of these findings, we further analyzed Olink data from the UK Biobank database, exploring the relationship between fasting duration and insulin receptor (INSR) expression, as well as the association between INSR expression levels and patient survival outcomes. Spearman rank correlation analysis revealed that longer fasting duration significantly reduced INSR expression (r = -0.15, P < 0.001) (Figure 4a). Mann-Whitney U tests showed significant differences in INSR expression between the fasting and satiety groups, with the fasting group exhibiting markedly lower INSR levels (P < 0.001) (Figure 4b). Further Kaplan-Meier survival analysis demonstrated that low INSR expression was associated with better survival outcomes (P = 0.0072) (Figure 4c). These analyses further confirm that fasting significantly modulates insulin levels, influences the insulin receptor signaling pathway, and plays an important role in improving survival outcomes for cancer patients.

**Figure 4:**
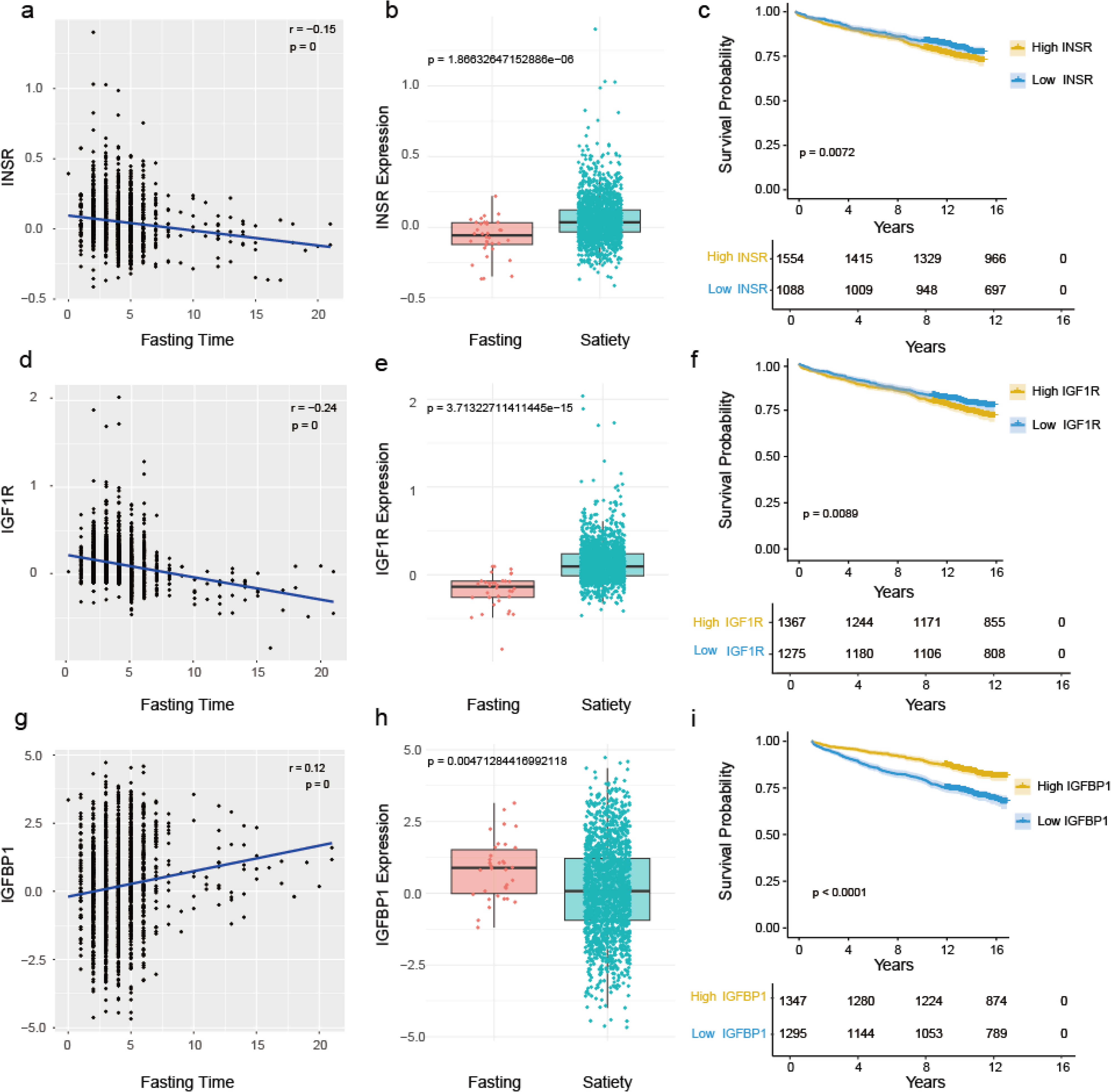
Correlation scatter plots, differential box plots, and survival curves of INSR, IGF1R, and IGF-BP1 from the UK Biobank database

Insulin-like growth factor 1 (IGF-1) is an important growth factor that plays a key role in cell proliferation, survival, and metabolic regulation (39). Recent studies have shown that IGF-1 is closely associated with the onset and progression of tumors. Four studies (27, 35, 37, 38) reported on the levels of IGF-1 under fasting intervention. Initial heterogeneity tests revealed significant variability across the four studies (*I²*=72%, P=0.01) (Supplementary Figure 1c). To further explore the source of heterogeneity, we conducted a sensitivity analysis and found that the study by Enas M. Omar et al. 2022 (35) had a substantial impact on the stability of the results (Supplementary Figure 2c). The analysis suggested that this could be due to differences in the fasting protocol used in this study compared to the others. After excluding this study, we conducted another heterogeneity test, which showed a significant reduction in heterogeneity (*I²*=50%, P=0.13) (Figure 3c). Based on this, we performed further data analysis using a fixed-effects model, and the results indicated that IGF-1 levels in the fasting group were significantly lower than those in the normal diet group (MD=-23.88, 95%CI [-41.41, -6.35], P=0.008). This suggests that fasting may inhibit tumor growth and spread to some extent by lowering IGF-1 levels, providing new theoretical support for fasting as an adjunctive treatment in cancer therapy. To further validate the potential significance of this finding, we used Olink data from the UK Biobank database to analyze the relationship between fasting duration and insulin-like growth factor 1 receptor (IGF1R) expression levels in tumor patients, as well as the association between IGF1R expression and patient survival prognosis. Spearman rank correlation analysis revealed that longer fasting duration significantly reduced IGF1R expression (r = -0.24, P < 0.001) (Figure 4d). Additionally, Mann-Whitney U tests showed significant differences in IGF1R expression between the fasting and satiety groups, with the fasting group exhibiting significantly lower IGF1R expression (P < 0.001) (Figure 4e). Further Kaplan-Meier survival analysis demonstrated that low IGF1R expression was significantly associated with better survival prognosis (P = 0.0089) (Figure 4f). These results further support the potential role of fasting in inhibiting tumor growth and improving overall survival by modulating IGF-1 and IGF1R levels.

Insulin-like growth factor-binding proteins (IGF-BPs) are natural inhibitors of IGF-1 and play a crucial role in various physiological processes, particularly in tumor microenvironments and the regulation of tumor cell metabolism (40). The levels of IGF-BPs are closely related to the occurrence, development, and treatment response of tumors. Therefore, we included three studies (27, 35, 37) to compare the levels of IGF-BP between the fasting group and the normal diet group. Initial heterogeneity tests showed significant heterogeneity between the studies (*I²*=63%, P=0.07) (Supplementary Figure 1d). To further explore the source of heterogeneity, we performed a sensitivity analysis, which indicated that the study by Stefanie de Groot et al. 2015(27), due to its smaller sample size, might have a large impact on the overall stability of the results (Supplementary Figure 2d). After excluding this study, no significant heterogeneity remained (*I²*=0, P=0.78) (Figure 3d). Thus, we performed further analysis using a fixed-effects model. The results showed that the IGF-BP levels in the fasting group were significantly higher than those in the normal diet group (MD=17.02, 95%CI [2.82, 31.21], P=0.02). This suggests that fasting may regulate IGF-BP levels, significantly affecting tumor cell proliferation and growth, thereby providing a positive adjunct to tumor therapy. To validate the clinical significance of this finding, we further analyzed Olink data from the UK Biobank database, exploring the relationship between fasting duration and the expression levels of insulin-like growth factor-binding protein 1 (IGFBP1), as well as the association between IGFBP1 expression and survival prognosis in cancer patients. Spearman rank correlation analysis showed that longer fasting duration significantly increased IGFBP1 expression (r = 0.12, P < 0.001) (Figure 4g). Moreover, Mann-Whitney U tests revealed significant differences in IGFBP1 expression levels between the fasting and satiety groups, with the fasting group exhibiting significantly higher IGFBP1 expression (P < 0.001) (Figure 4h). Further Kaplan-Meier survival analysis showed that high IGFBP1 expression was significantly associated with better survival prognosis (P < 0.001) (Figure 4i). The Olink data further support that fasting, by regulating IGF-BP levels, may positively impact tumor growth and is associated with better survival outcomes. Overall, the potential of fasting as an adjunctive therapy in cancer treatment warrants further investigation and exploration.

### 3.4 Fasting doesn’t increase treatment-related toxicity in cancer patients

The main therapeutic approaches for cancer remain chemotherapy, radiotherapy, and immunotherapy. While these treatments can significantly extend survival, improve quality of life, and effectively control tumor progression, they inevitably lead to adverse events and hematological toxicity, which are crucial concerns during clinical treatment (41). To assess the impact of fasting on adverse events in cancer patients, we included five relevant studies (26, 27, 32, 35, 38) to analyze the incidence of total grade III/IV adverse events in fasting versus normal diet groups. Initial heterogeneity tests revealed no significant differences between the studies (*I²*=0, P=0.51) (Supplementary Figure 3a). Fixed-effects model analysis showed that the incidence of t total grade III/IV adverse events was not significantly different between the fasting and normal diet groups (RR=1.13, 95% CI [0.91, 1.42], P=0.27). To further enhance the reliability of the analysis, we specifically examined the incidence of two common adverse events: grade III/IV neutropenia and grade III/IV neutropenic fever. Heterogeneity tests for these two adverse events also showed no significant differences between the studies (*I²*=0, P=0.42; *I²*=0, P=0.4) (Supplementary Figure 3b and 3c). The fixed-effects model analysis again indicated no significant differences in the incidence of these two adverse events between the fasting and normal diet groups (RR=1.34, 95% CI [0.89, 2.01], P=0.16; RR=1.66, 95% CI [0.94, 2.94], P=0.08). This study demonstrates that fasting does not significantly increase the incidence of grade III/IV adverse events in cancer patients. It further confirms the safety and tolerability of fasting as an adjunct in cancer treatment, providing new insights for optimizing and expanding future cancer treatment strategies.

Changes in blood component levels are often used as important indicators to evaluate the impact of cancer treatments on the hematological system. When exploring the potential benefits of fasting for cancer patients, it is crucial to assess whether fasting might increase hematological toxicity to determine its safety (42). To further investigate the effect of fasting on hematological safety, we first analyzed the impact of fasting on erythrocyte count by including two relevant studies (35, 38). Heterogeneity testing of the two included studies showed no significant heterogeneity between them (*I²*=0, P=0.84) (Supplementary Figure 4a). Based on this, a fixed-effects model was used for data analysis, which revealed no significant difference in erythrocyte count between the fasting and normal diet groups (MD=0.03, 95% CI [-0.11, 0.17], P=0.64). This finding further confirms that fasting does not negatively affect erythrocyte count, suggesting that fasting is safe for the hematological system and helps maintain blood component stability. Additionally, hemoglobin (HG), a key protein in red blood cells, plays an important role in hematological safety. To further verify the impact of fasting on erythrocyte function, we also analyzed its potential effect on hemoglobin levels. Three studies (32, 35, 38) were included in this analysis, and heterogeneity testing revealed no significant differences (*I²*=0, P=0.95) (Supplementary Figure 4d). Fixed-effects model analysis also showed no significant difference in hemoglobin levels between the fasting and normal diet groups (MD=-0.14, 95% CI [-0.50, 0.23], P=0.47). This finding further supports the idea that fasting has only a minimal effect on erythrocytes and related proteins, particularly hemoglobin, reinforcing the safety of fasting as an adjunct in cancer treatment. To validate the clinical significance of these findings, we analyzed Olink data from the UK Biobank to explore the relationship between fasting duration and the expression levels of Transferrin (TF), Hemoglobin subunit theta-1 (HBQ1), and Hemoglobin subunit zeta (HBZ), and how these levels relate to patient prognosis. Transferrin (TF) is responsible for safely transporting iron to cells and is closely related to iron metabolism and erythrocyte production, while HBQ1 and HBZ, as subunits of hemoglobin, indirectly reflect hemoglobin levels. Spearman rank correlation analysis showed no significant correlation between fasting duration and TF, HBQ1, and HBZ expression levels (r=0.04, P=0.095; r=0.04, P=0.12; r=0.01, P=0.65) (Supplementary Figure 5a, 5d and 5g). Further Mann-Whitney U tests showed no significant differences between the fasting and satiety groups in TF, HBQ1, and HBZ expression levels (P=0.31; P=0.27; P=0.43) (Supplementary Figure 5b, 5e and 5h). Additionally, Kaplan-Meier survival analysis indicated that the expression levels of TF, HBQ1, and HBZ did not significantly affect cancer patient survival prognosis (P=0.15; P=0.96; P=0.81) (Supplementary Figure 5c, 5f and 5i).

Next, we included two studies (35, 38) to investigate the effect of fasting on leukocyte levels. The heterogeneity test results showed no significant heterogeneity between the two studies (*I²*=0, P=0.52) (Supplementary Figure 4b). Based on this, we performed the analysis using a fixed-effects model, and the results indicated no significant difference in leukocyte levels between the fasting and normal diet groups (MD=-0.43, 95% CI [-1.10, 0.24], P=0.21). This further suggests that fasting does not significantly affect leukocyte levels and does not pose a risk of altering leukocyte counts, supporting the idea that fasting helps maintain the stability of the immune system. As neutrophils are one of the most important cell types within the leukocyte category, we further analyzed the effect of fasting on neutrophil levels (32, 35, 38). The heterogeneity test results showed no significant heterogeneity between these studies either (*I²*=0, P=0.7) (Supplementary Figure 4e). The fixed-effects model analysis revealed no significant difference in neutrophil levels between the fasting and normal diet groups (MD=-0.31, 95% CI [-0.88, 0.27], P=0.3). This result is consistent with the leukocyte analysis, demonstrating that fasting helps maintain the stability of immune components in the blood, thereby supporting normal immune function. To further verify the clinical significance of these results, we analyzed Olink data from the UK Biobank to explore the relationship between fasting duration and the expression levels of neutrophil-related biomarkers (such as Granulocyte colony-stimulating factor [CSF3], Interleukin-8 [CXCL8], Myeloperoxidase [MPO], and Tumor Necrosis Factor-alpha [TNF]), and their association with tumor patient survival outcomes. Spearman rank correlation analysis showed no significant correlation between fasting duration and the expression of CSF3 and MPO (r=-0.02, P=0.49; r=0, P=0.83) (Supplementary Figure 7a and 7g). However, fasting duration showed a weak but significant negative correlation with the expression levels of CXCL8 and TNF (r=-0.06, P=0.02; r=-0.06, P=0.006) (Supplementary Figure 7d and 7j).Further Mann-Whitney U test results indicated no significant differences in the expression levels of CSF3, CXCL8, MPO, and TNF between the fasting and fed groups (P=0.95; P=0.21; P=0.29; P=0.09) (Supplementary Figure 7b, 7e, 7h, and 7k). Additionally, Kaplan-Meier survival analysis showed no significant impact of these biomarkers’ expression levels on tumor patient prognosis (P=0.21; P=0.62; P=0.07; P=0.06) (Supplementary Figure 7c, 7f, 7i, and 7l).

To comprehensively assess the impact of fasting on the hematological safety of cancer patients, we further analyzed the potential effect of fasting on platelet levels. Three relevant studies (32, 35, 38) were included. After performing heterogeneity tests, no significant heterogeneity was found (*I²* = 0, P = 0.77) (Supplementary Figure 4c). Based on this, we used a fixed-effect model for analysis, and the results showed no significant difference in platelet levels between the fasting group and the normal diet group (MD = 18.81, 95% CI [-5.08, 42.69], P = 0.12). We analyzed data from the UK Biobank using Olink data to explore the relationship between fasting duration and the expression levels of two subunits of Platelet glycoprotein Ib (Platelet glycoprotein Ib alpha chain [GP1BA] and Platelet glycoprotein Ib beta chain [GP1BB]), and examined the association between GP1BA and GP1BB expression and survival prognosis in cancer patients. Spearman rank correlation analysis showed a statistically significant but weak negative correlation between fasting duration and GP1BA and GP1BB expression levels (r = -0.08, P < 0.001; r = -0.10, P < 0.001) (Supplementary Figure 6a and 6d). Further Mann-Whitney U tests indicated no significant difference in GP1BA and GP1BB expression levels between the fasting group and the fed group (P = 0.11; P = 0.08) (Supplementary Figure 6b and 6e). Moreover, Kaplan-Meier survival analysis showed no significant impact of GP1BA and GP1BB expression levels on the survival prognosis of cancer patients (P = 0.69; P = 0.22) (Supplementary Figure 6c and 6f).Overall, the analysis indicates that fasting does not have a significant effect on hematological toxicity in cancer patients, further confirming the safety of fasting in cancer treatment, and demonstrates its ability to maintain hematological stability, providing support for its application in cancer therapy.

### 3.5 Fasting has no significant impact on the scale scores of cancer patients

In cancer treatment, patients’ health-related quality of life (QOL) and the degree of treatment-induced distress are important indicators for assessing treatment efficacy and optimizing therapeutic strategies(43). To explore the impact of fasting on the quality of life in cancer patients, several studies have used different scales for evaluation, including the FACIT© system, the global QoL scale, and the distress thermometer. Stephan P. Bauersfeld et al. 2018(28) used the FACIT© system to divide patients into two groups, A and B. Group A underwent STF intervention during the first three cycles and normal diet during the last three cycles, while Group B followed the opposite schedule. The study found significant differences in FACT-G, FACIT-F, TOI-FACIT-F, and total FACIT-F scores between fasting and normal diet periods (C1-C3 vs. C4-C6) in Group A, indicating that STF intervention significantly improved QOL and fatigue. However, in Group B, no significant changes were observed in QOL and fatigue scores during chemotherapy, suggesting that the impact of fasting on quality of life may vary depending on the treatment context. The discrepancy between Groups A and B could be related to differences in the ovarian cancer treatment protocols of the two groups. Stefanie de Groot et al. 2020(26) and Rieneke T. Lugtenberg et al. 2021(29) used the global QoL scale and distress thermometer, respectively, to assess patients’ health status and distress levels. The results showed a significant decline in global QoL scores during chemotherapy, which then returned to baseline levels during follow-up. Although distress scores increased gradually during treatment and follow-up, no significant differences were found between the two groups. Overall, these studies suggest that while quality of life is generally affected during treatment, the degree of distress and the recovery of quality of life did not show significant differences between the groups.

### 3.6 Publication bias

In the outcome indicators mentioned above, only the overall incidence of grade III/IV adverse effects included ≥5 studies. A funnel plot for the total grade III/IV adverse effects was generated using Stata 18 (Supplementary Figure 8). Additionally, Begg’s test for publication bias yielded P=1.00 > 0.05, and Egger’s test resulted in P=0.768 > 0.05, indicating that there is no publication bias among the five studies included in the total grade III/IV adverse effects. Furthermore, among the above indicators, there are six outcomes with more than two studies included, namely: grade III/IV neutropenia, insulin, IGF-1, platelets, hemoglobin, and neutrophils. Egger’s test for publication bias was performed on these six outcomes using Stata 18, and it was found that except for grade III/IV neutropenia, the P-values for the other five outcomes were all greater than 0.05, indicating no publication bias for these outcomes. However, for grade III/IV neutropenia, the Egger’s test P-value was 0.007 < 0.05, suggesting potential publication bias, which requires further assessment using the trim-and-fill method (Supplementary Figure 9). The original pooled effect size was RR=1.34 (95% CI: 0.89-2.01), which was not statistically significant. After adjusting for potential missing studies using the trim-and-fill method, the pooled effect size increased to RR=1.477 (95% CI: 0.989-2.205), but it still did not reach statistical significance. These results suggest that while there may be slight publication bias in the studies included, its impact on the incidence of grade III/IV neutropenia is limited.

## 4. Discussion

The impact of fasting on the treatment efficacy of cancer patients has recently become a hot topic (44). Given the potential risks associated with fasting, it is important to evaluate how different fasting protocols affect various efficacy outcomes and treatment-related toxicity across different types of cancer. In our study, we found that fasting significantly reduces metabolic parameters such as glucose, insulin, and IGF-1, while significantly increasing IGF-BP levels. Additionally, fasting reducing DNA damage in CD45+CD14+CD15-monocytes and increase the rates of complete response (CR) and partial response (PR). Importantly, there were no significant differences observed in terms of adverse effects, hematologic toxicity, or subjective assessments of patient physical status as measured by various scales. Although the changes in outcome indicators between fasting and control groups in the included studies may not align with the results of this analysis, the pooled analysis of outcome changes provides a more robust estimate.

To further explore the specific mechanisms underlying this effect, we reviewed a large body of basic research literature. Fasting can lower circulating glucose levels, reduce insulin levels, and decrease tumor PI3K signaling (45). Many studies have pointed out that the insulin receptor (INSR) is a type II receptor tyrosine kinase (RTK) involved in metabolism, growth, and proliferation (46). The expression of INSR is regulated by cell type, developmental stage, and disease state. In highly proliferative T cells, INSR expression increases, and INSR deficiency leads to impaired proliferation and immune dysfunction(47). Studies in mice have shown that glucose, as an inducer of the PI3K-Akt-mTOR pathway, plays a significant role in carcinogenesis. In tumor cells, the IGF-1 signaling pathway is often activated, causing tumor cells to direct more metabolic products and energy toward tumor growth and metastasis. The IGF-1-IGF-1R signaling pathway regulates various cellular phenotypes related to tumor cell survival and growth, including cell cycle progression, apoptosis, and differentiation(48, 49). Other cancer-related phenotypes regulated by IGF-1R signaling include cell adhesion and migration (50), as well as cancer metastasis(51, 52). IGFBPs play a central role in modulating the carcinogenic effects of IGF1R signaling because their affinity for IGF-1 and IGF-2 is typically greater than the affinity of IGF for IGF1R. Therefore, IGF bound to IGFBPs may be limited in activating IGF1R. Fasting reduces IGF-1 levels while increasing IGF-BP levels, thereby inhibiting signaling pathways and slowing tumor progression. These highly consistent with the findings of this study, fasting affects tumor treatment efficacy-related signaling pathways by altering glucose, insulin, IGF-1, and IGF-BP levels, thereby improving treatment outcomes. Furthermore, changes in insulin, IGF-1, and IGF-BP levels also contribute to prolonged overall survival (OS) in patients, positively impacting their prognosis.

The tumor treatment toxicities, represented by adverse effects and hematological toxicity, did not show significant changes due to fasting interventions. While protein expression in the blood can indirectly reflect hematological parameters, we found that these proteins did not appear to benefit patient prognosis. Although the quality of the included studies was relatively high, only a few clinical studies have evaluated the potential of different fasting regimens (e.g., STF, FMD, and IF) in improving tumor treatment toxicities. Therefore, whether fasting has a significant impact on toxicity still needs further investigation. All these clinical studies suggest that fasting, as an adjunctive therapy for cancer treatment, is safe and may have good tolerability, but it is not suitable for all patients.

This study also has some limitations. First, there is considerable variation in the sample sizes of the included studies, so the results should be interpreted with caution. Additionally, individual factors such as age, comorbidities, and medication treatments were difficult to consistently control across studies. Second, the exclusion of non-English published studies may introduce language bias, affecting the generalizability of the conclusions. Nutritional intervention research is inherently challenging, especially when evaluating and intervening in patients’ lifestyles, which can be influenced by cancer diagnosis. Furthermore, most studies relied on self-reported data, which may result in inaccurate information. Participant compliance with fasting and dietary interventions was generally low, especially during chemotherapy, which affected the assessment of intervention efficacy. Enhancing patient education, understanding, and engagement may play a key role in improving fasting adherence. Understanding the facilitators or barriers to patient compliance with fasting could improve adherence and clinical outcomes (53).

## 5. Conclusion

Fasting improved cancer treatment by reducing DNA damage in immune cells and altering the levels of various metabolites. Furthermore, we identified glucose, insulin, and IGF-1/IGF-BP levels as potential biomarkers for prognosis in cancer patients. Importantly, fasting does not induce additional treatment-related toxicities, providing strong support for its use as an adjunctive therapy in cancer treatment.

## 7. Author Contributions

Zeyao Wang: Contributed to the conceptualization of the study, data synthesis, and analysis. Also involved in drafting and revising the manuscript.Xueying Wang(Co-first Authors): Assisted in the study design, performed data analysis, and contributed to manuscript writing and revisions.Lina Cui(Co-first Authors): Contributed to data analysis, interpretation of results, and manuscript drafting.Zihan Liu: Supported the literature review, data synthesis, and interpretation of findings.Fanxuan Huang: Assisted with the data analysis and provided critical input during manuscript writing.Zhaoyu Pan: Contributed to the interpretation of results and provided relevant literature for analysis.Jiaqing Xiao: Conducted literature review, assisted with manuscript preparation, and provided revisions.Tong Liu(Corresponding author): Oversaw the study’s conceptualization, supervised the overall analysis, and critically revised the manuscript.

## 8. Data Share Statement

Data described in the manuscrint, code book, and analvtic code will be made available upon requestpending[e.g., application and approval, payment, other]

---

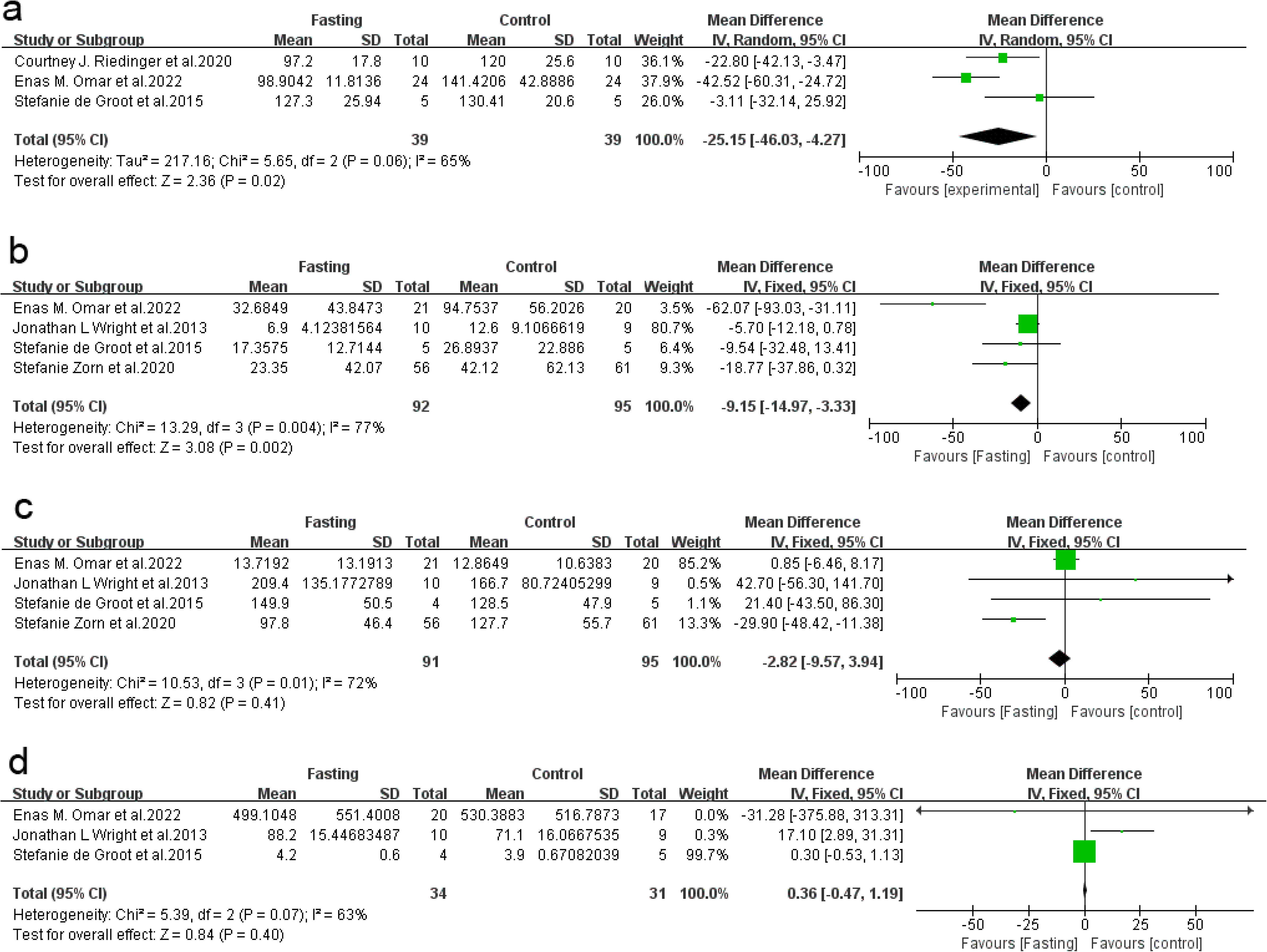

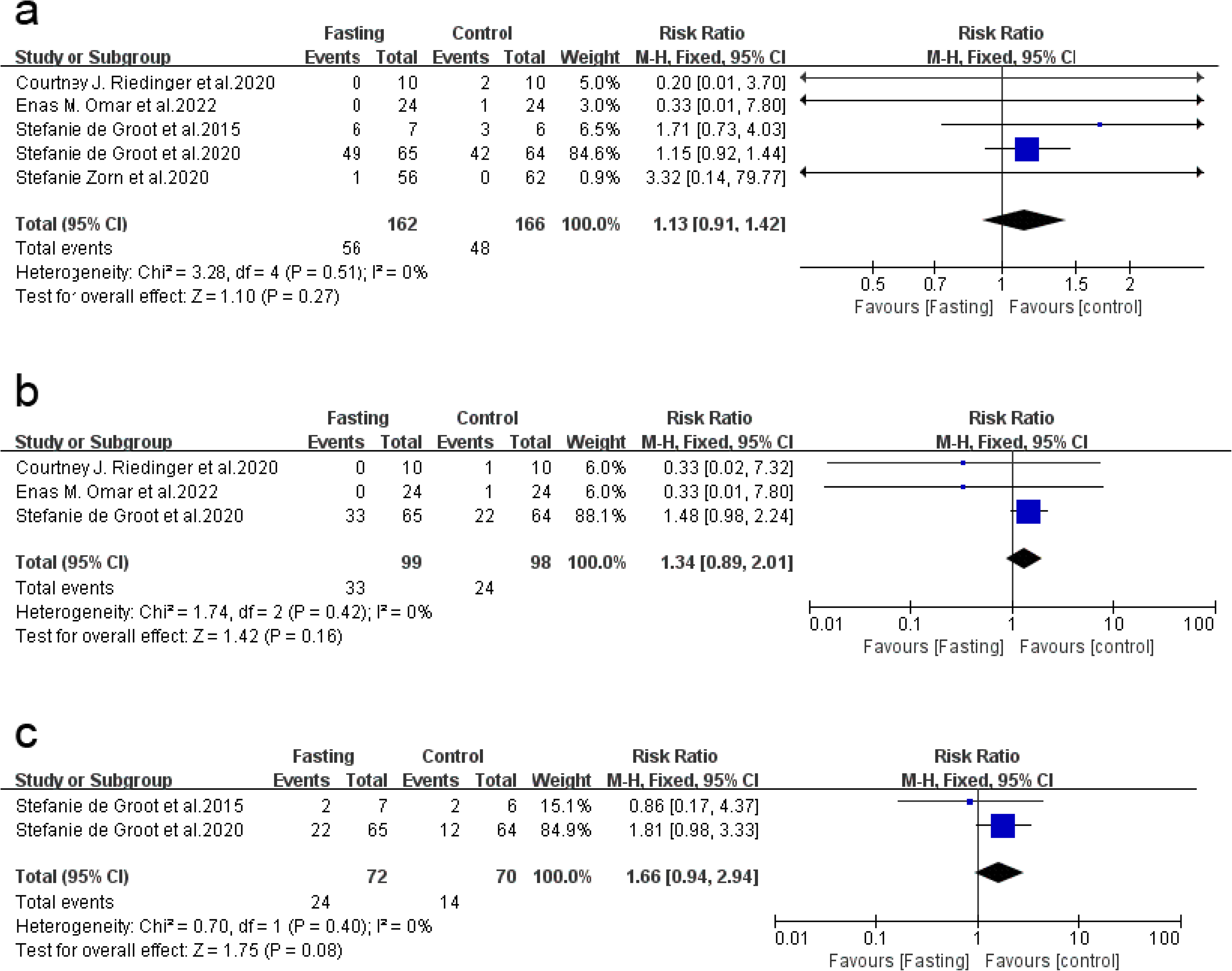

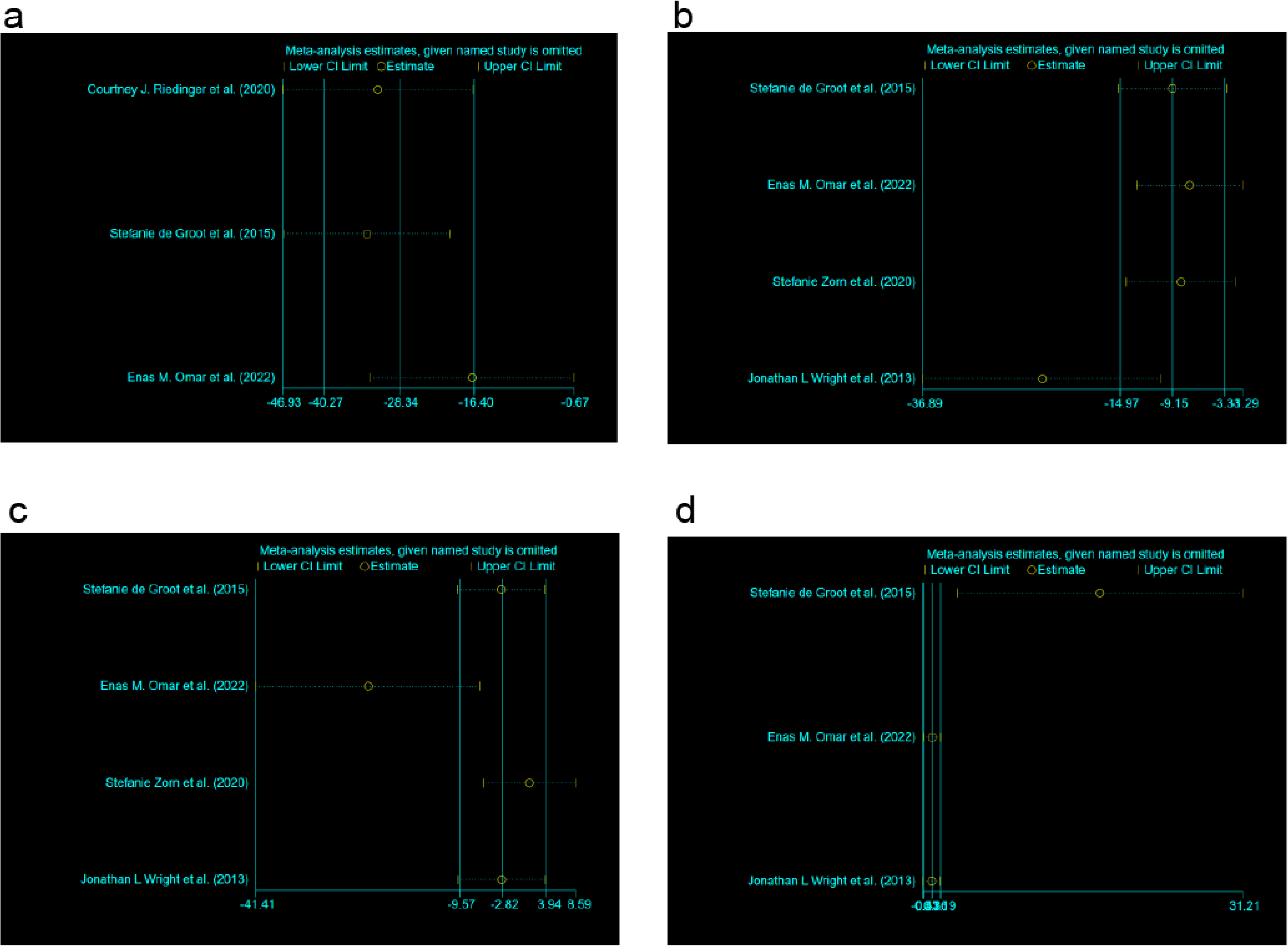

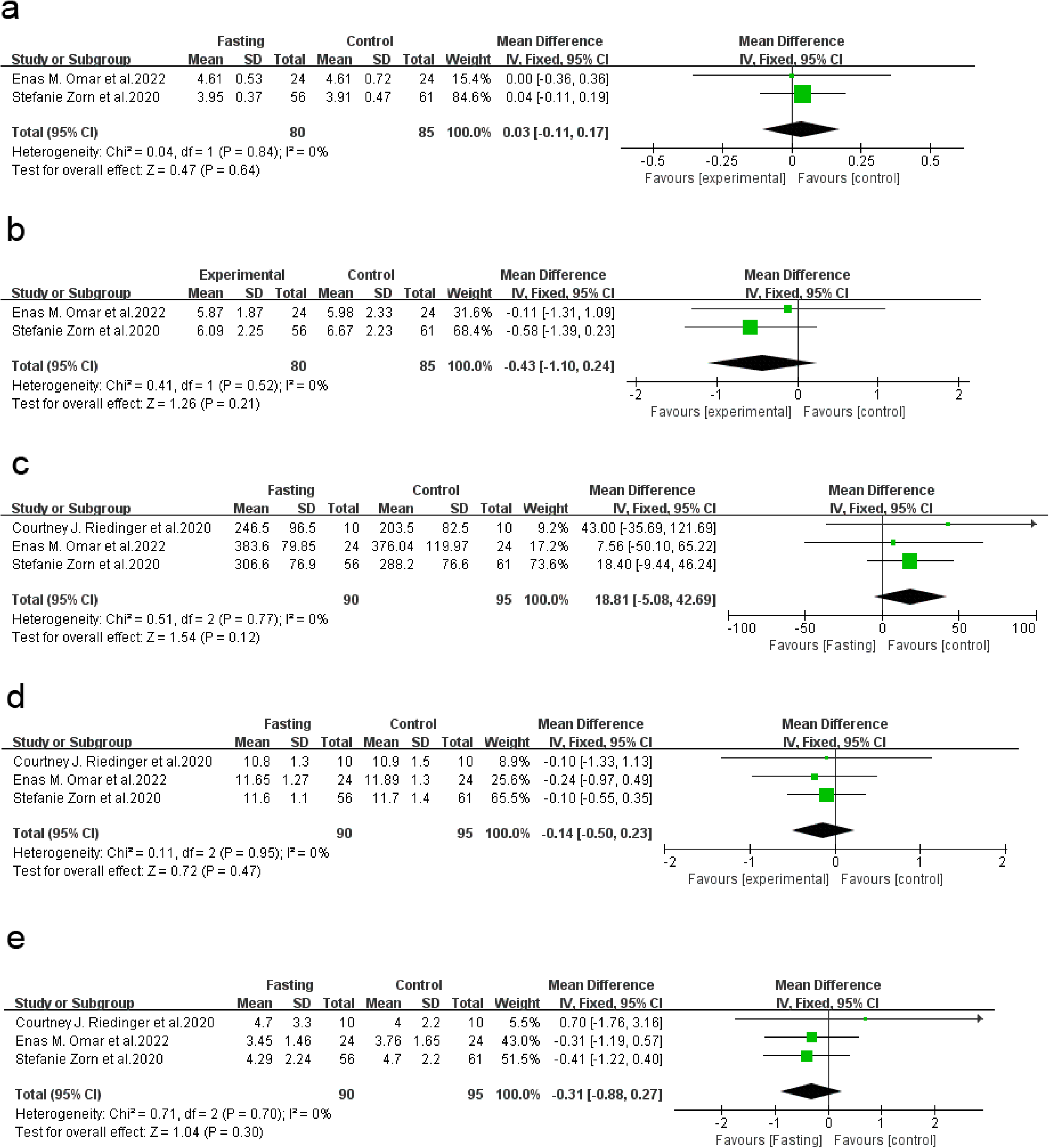

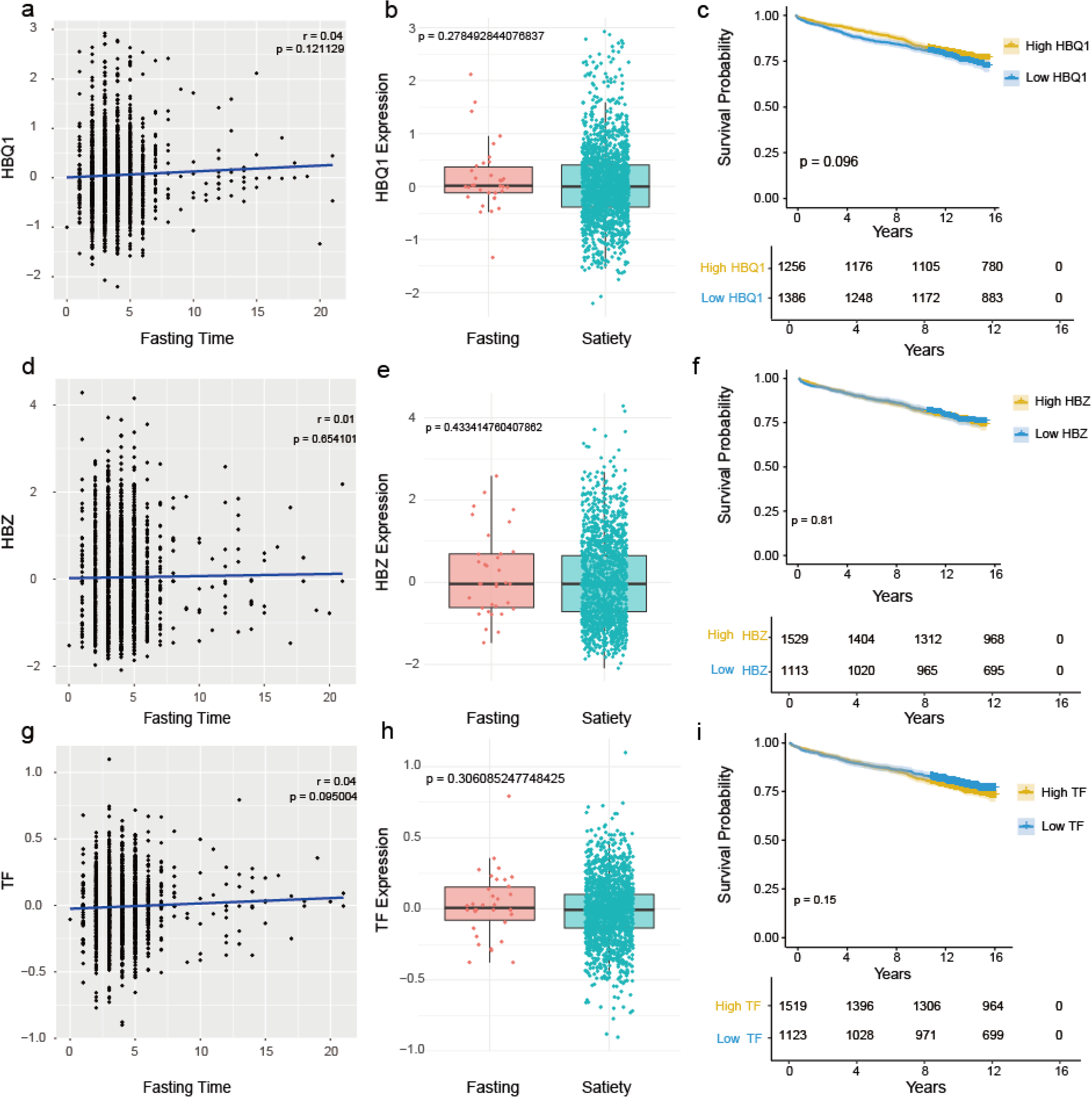

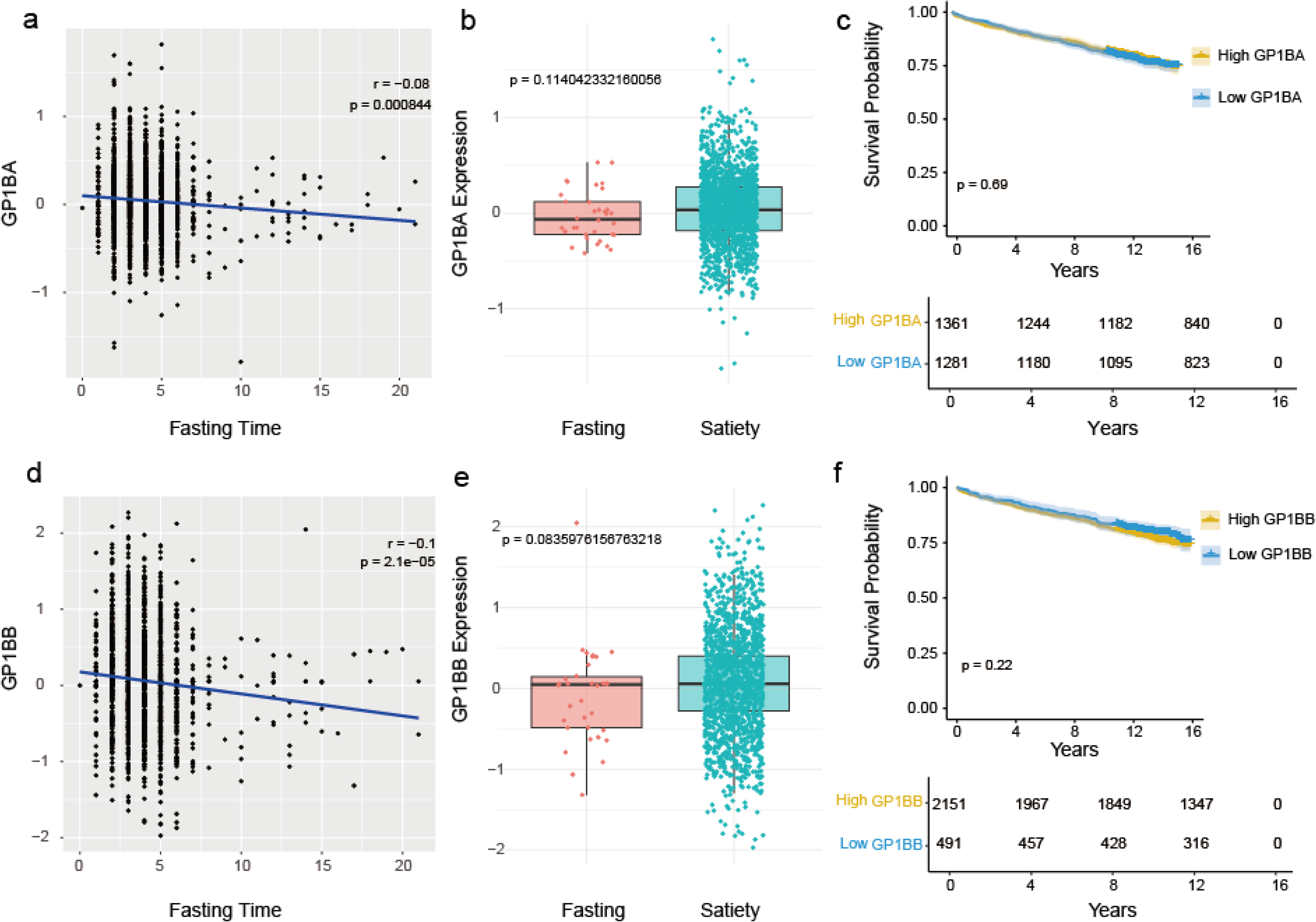

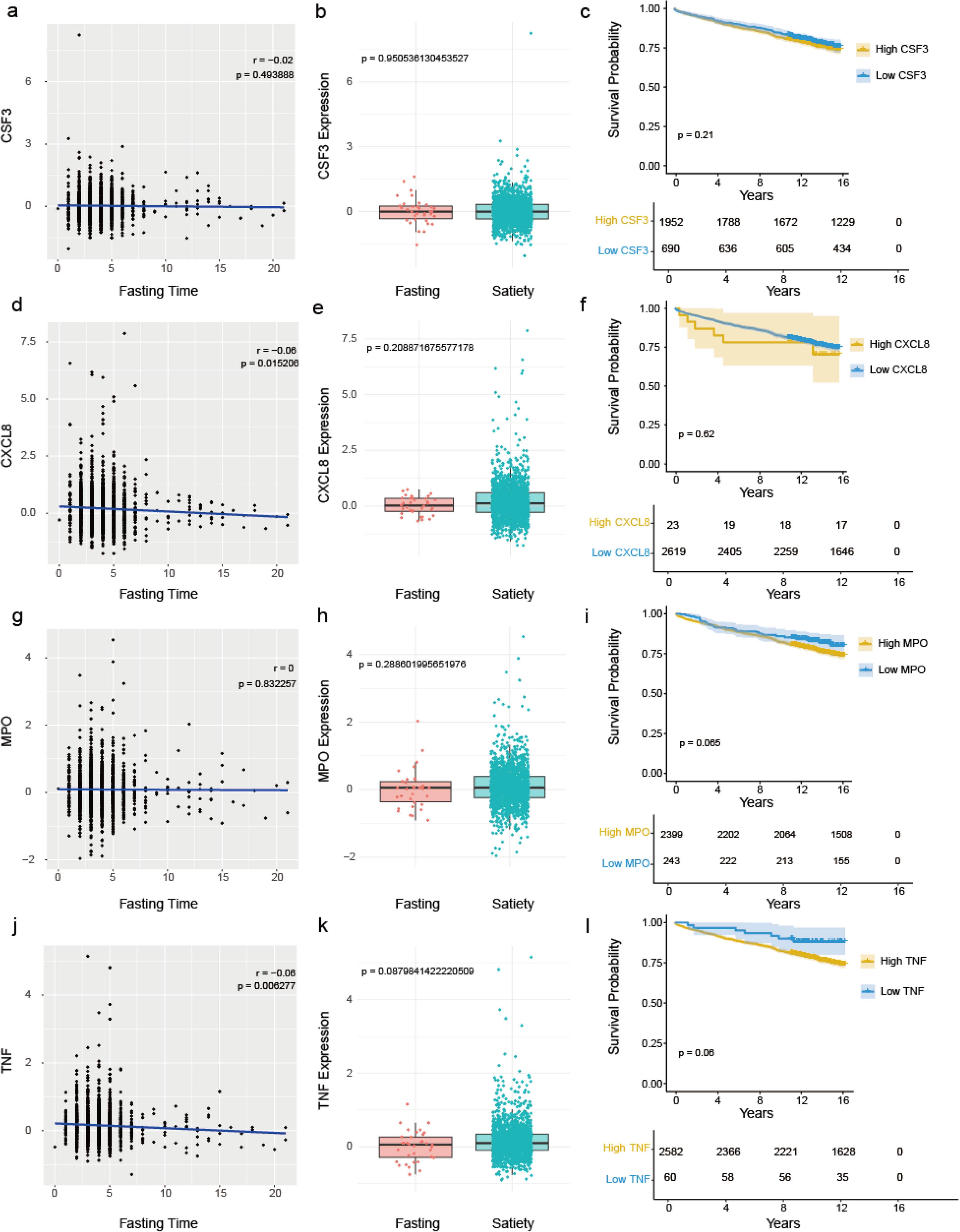

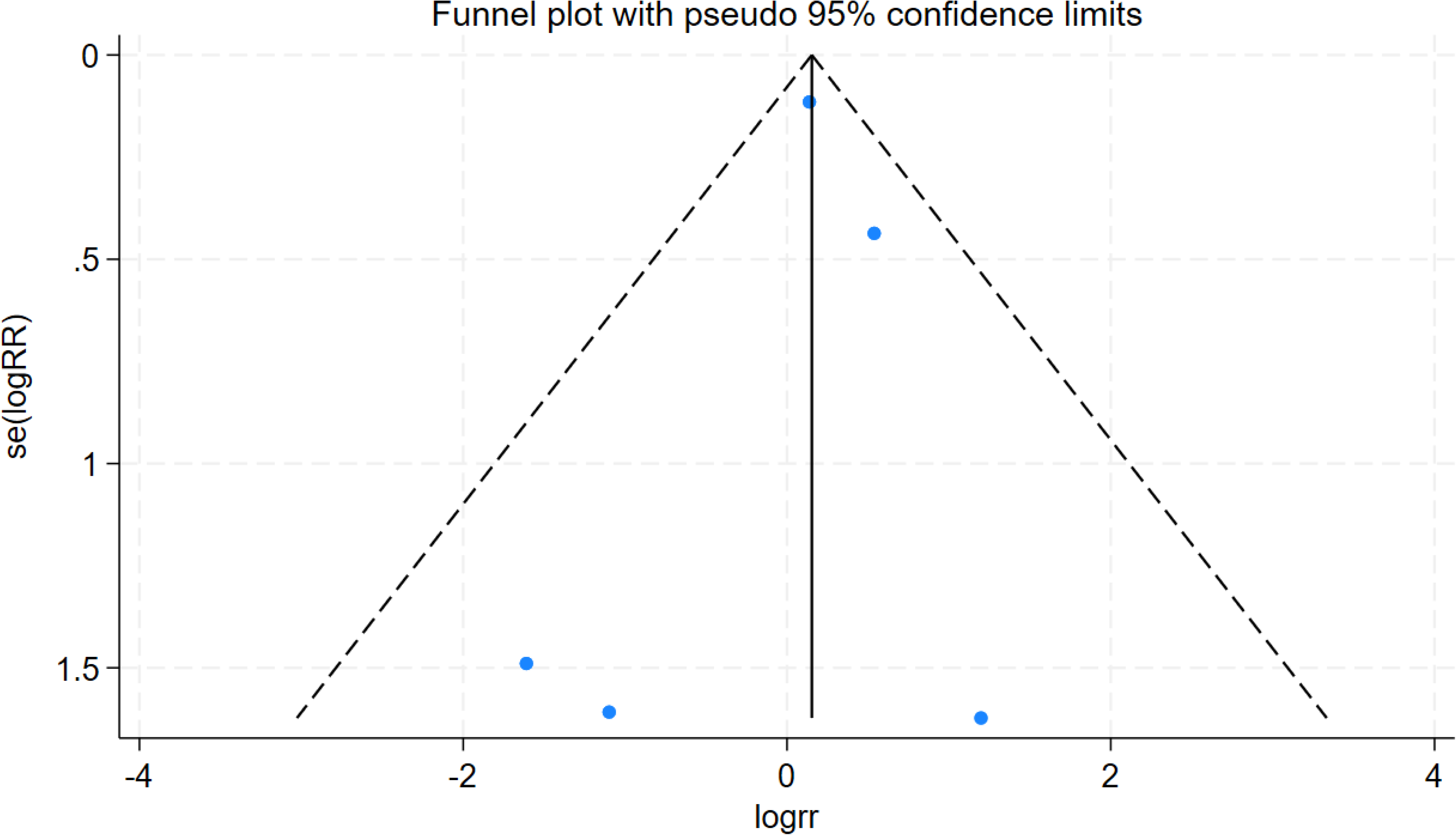

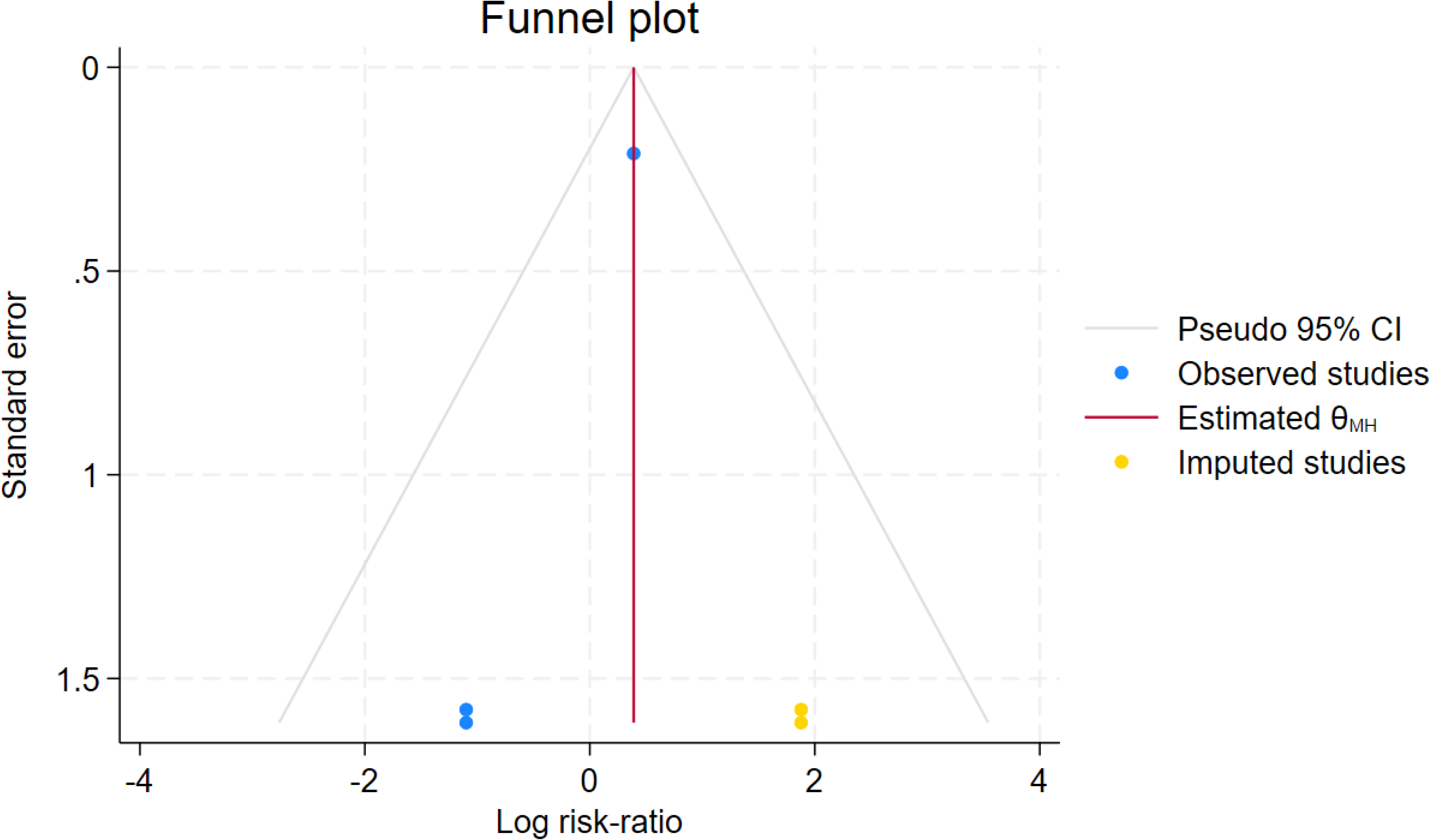

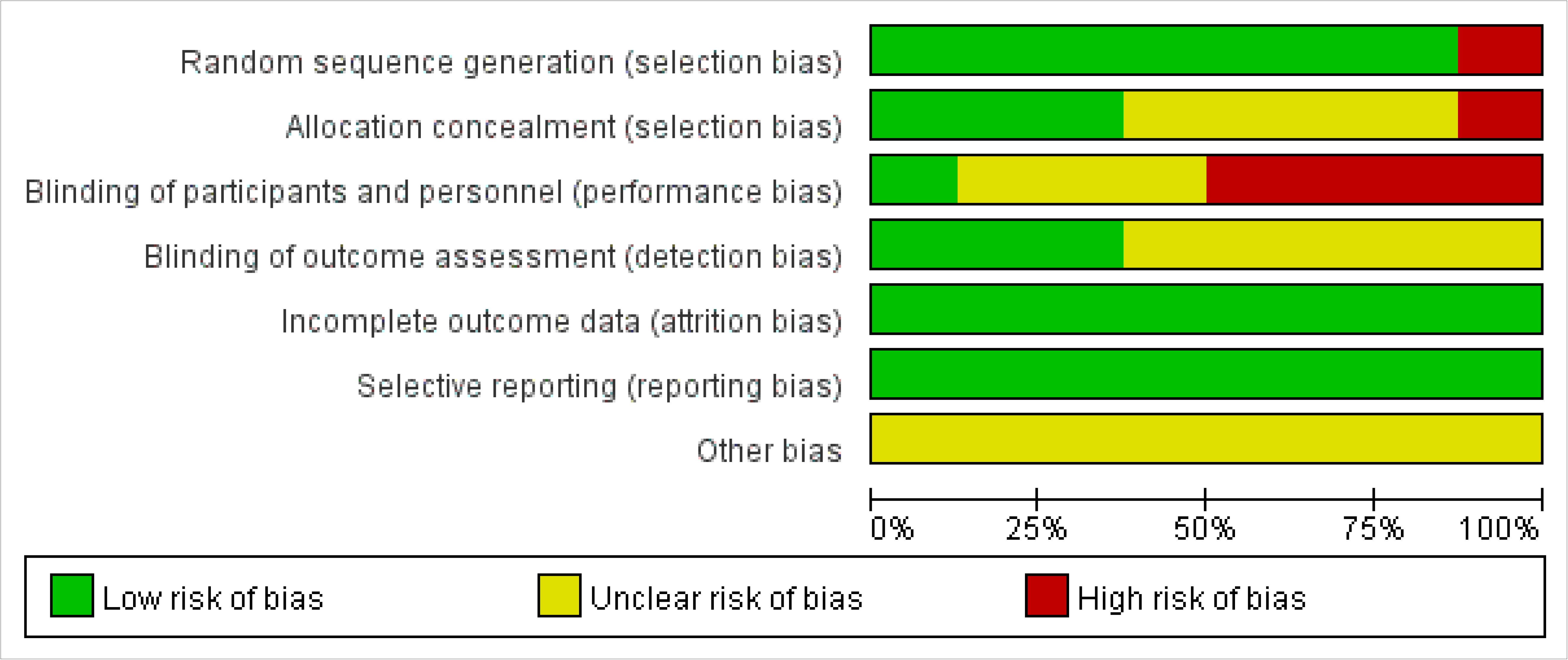

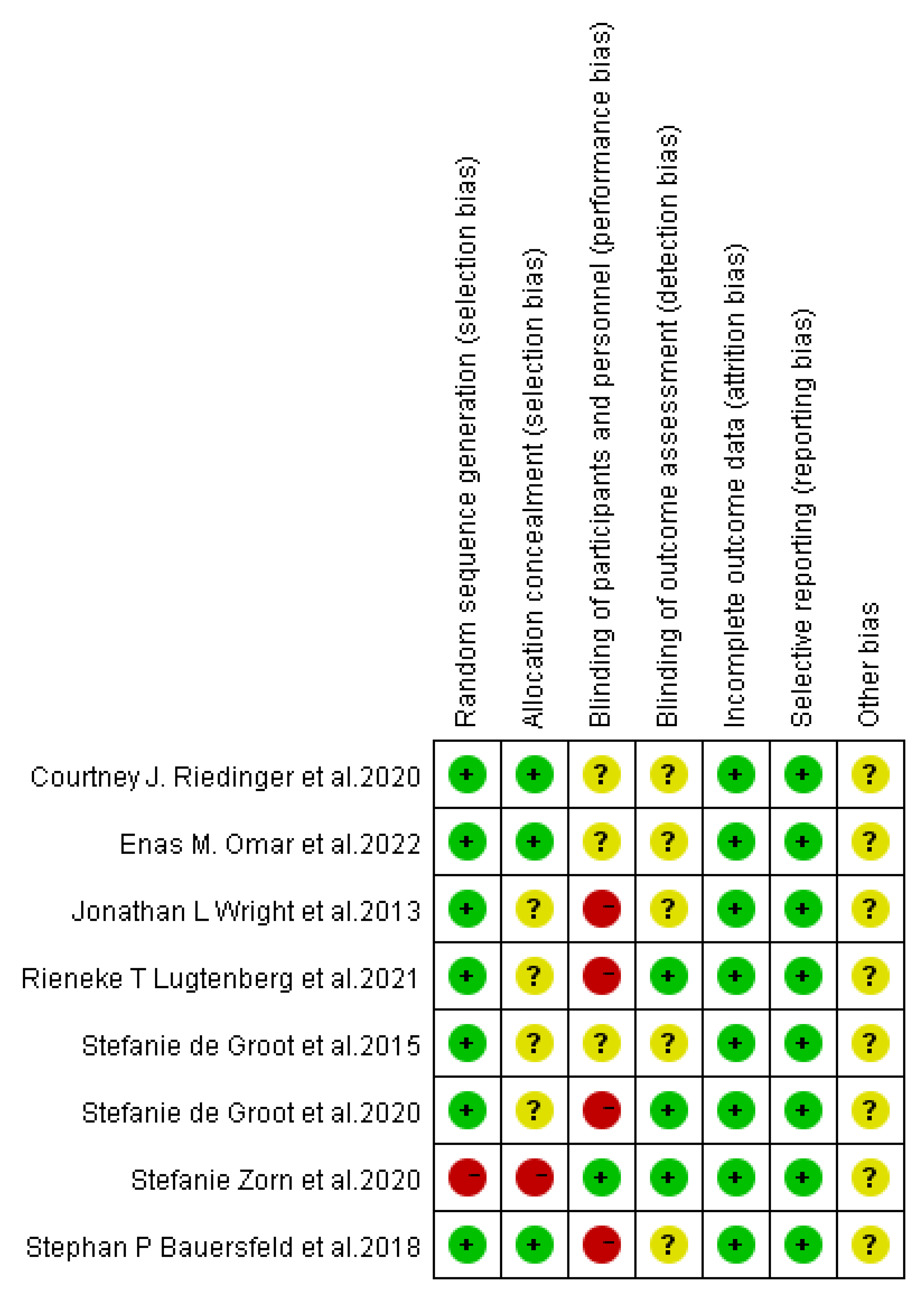

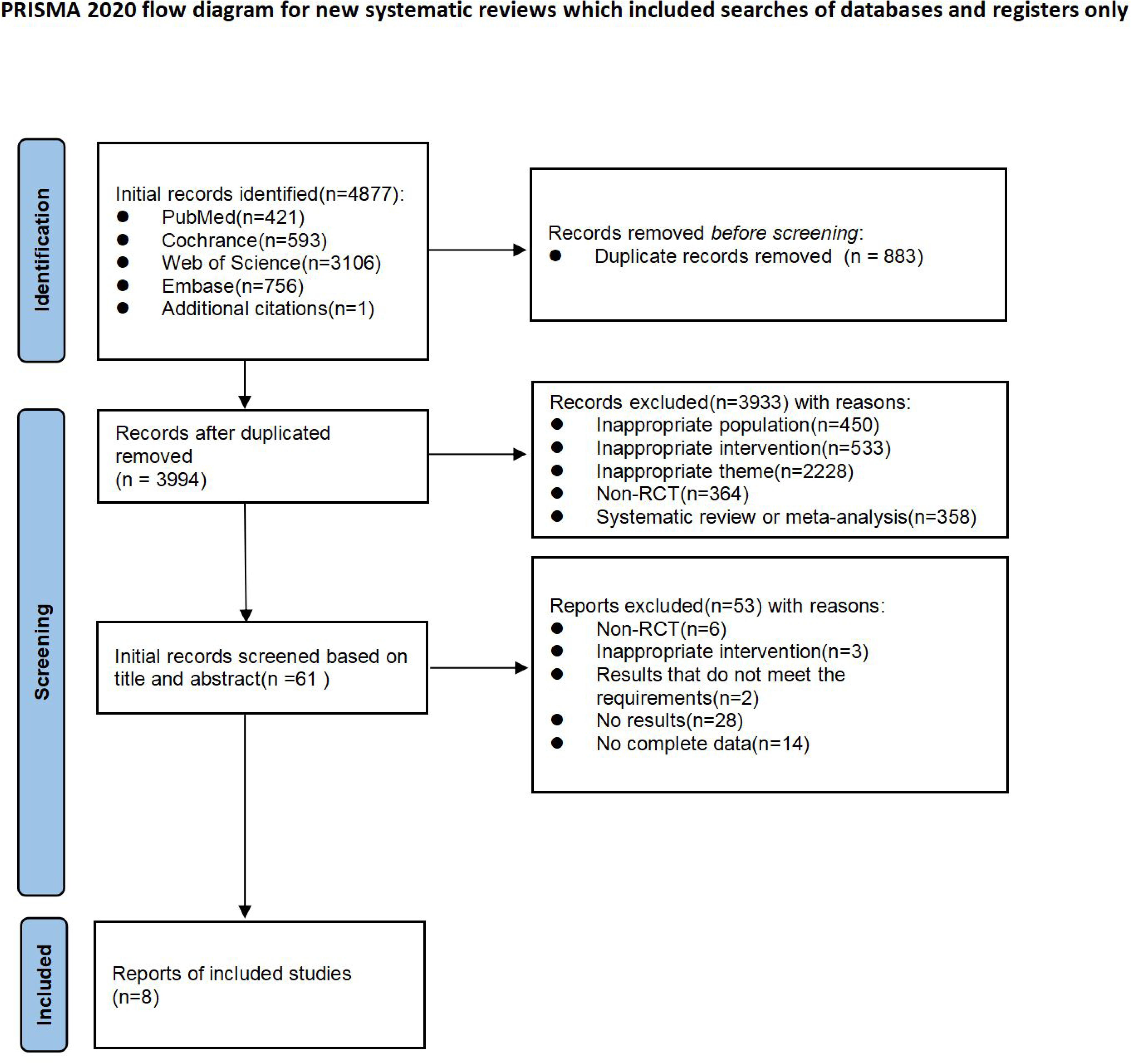

**Table S1:**
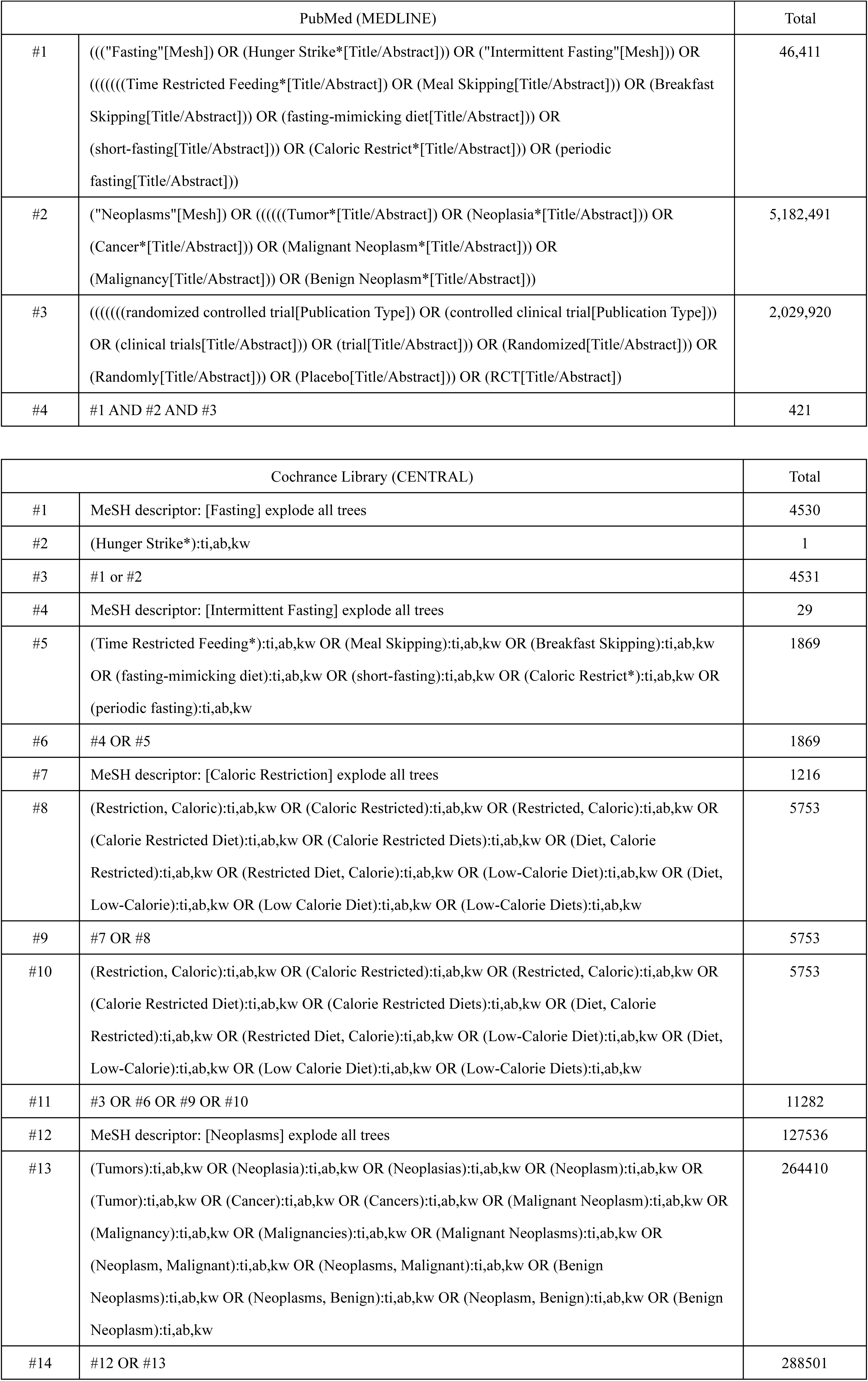
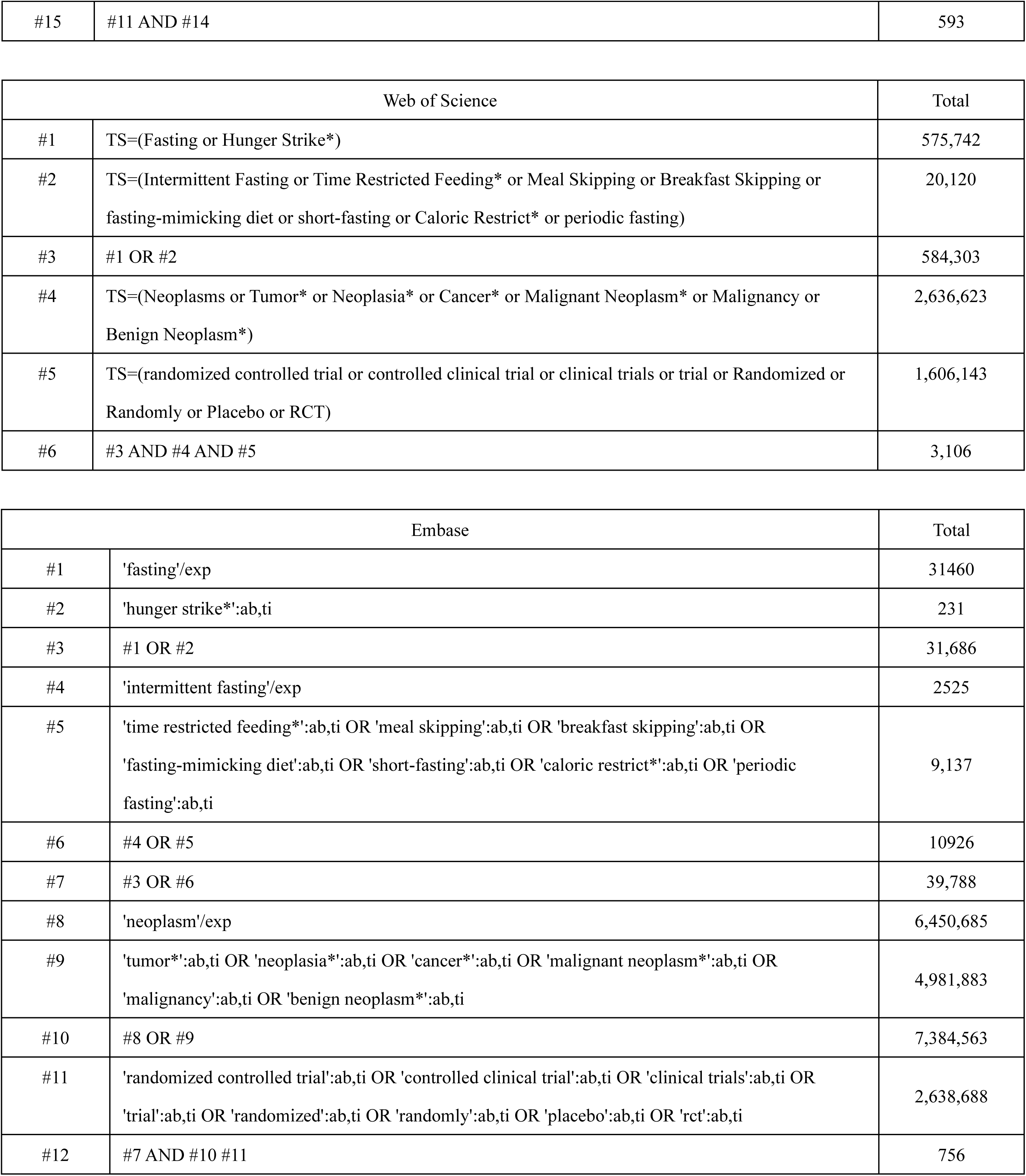
Search strategies for PubMed (MEDLINE), Web of Science, Embase, and Cochrane Library (CENTRAL)

**Table S2:**
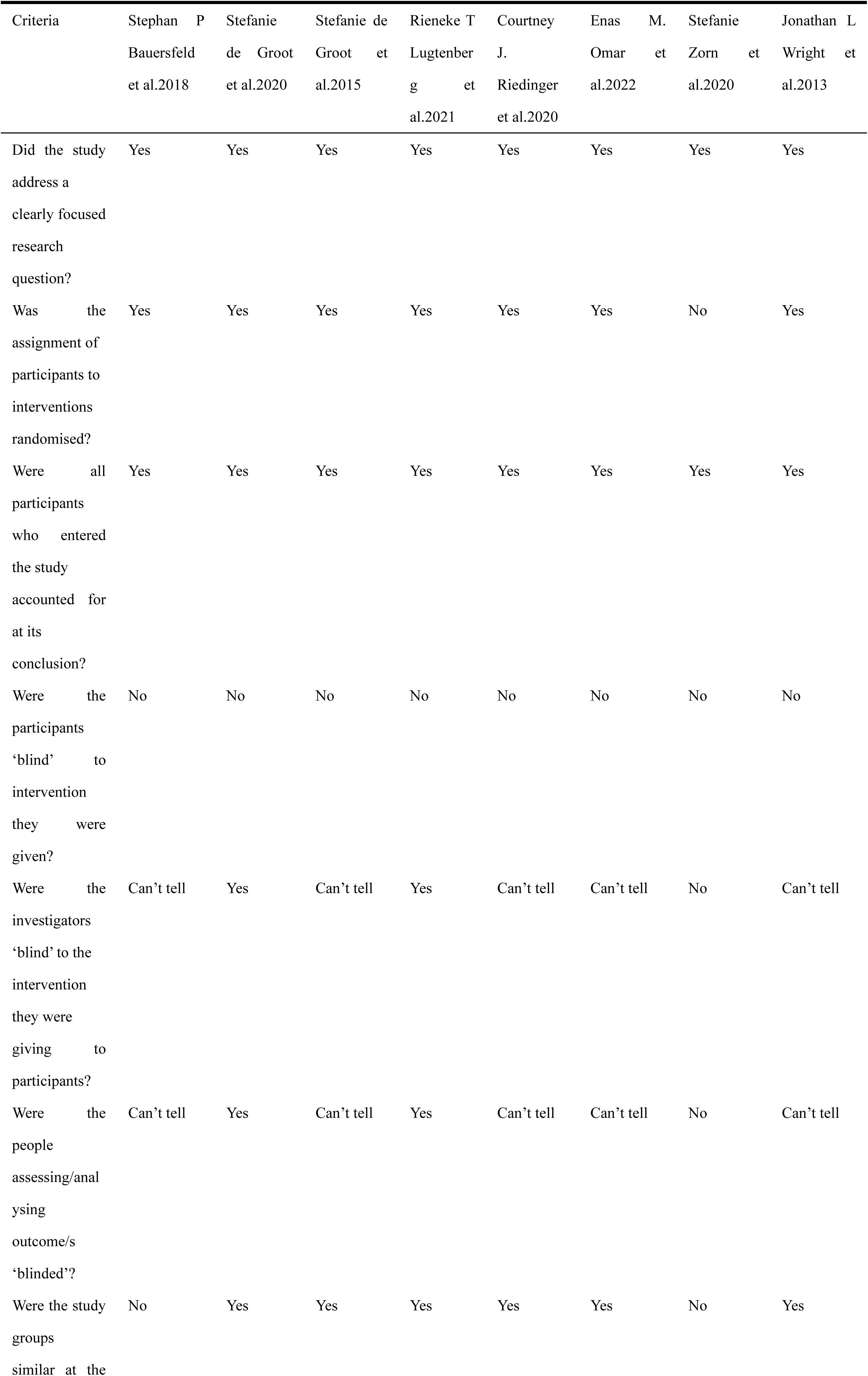
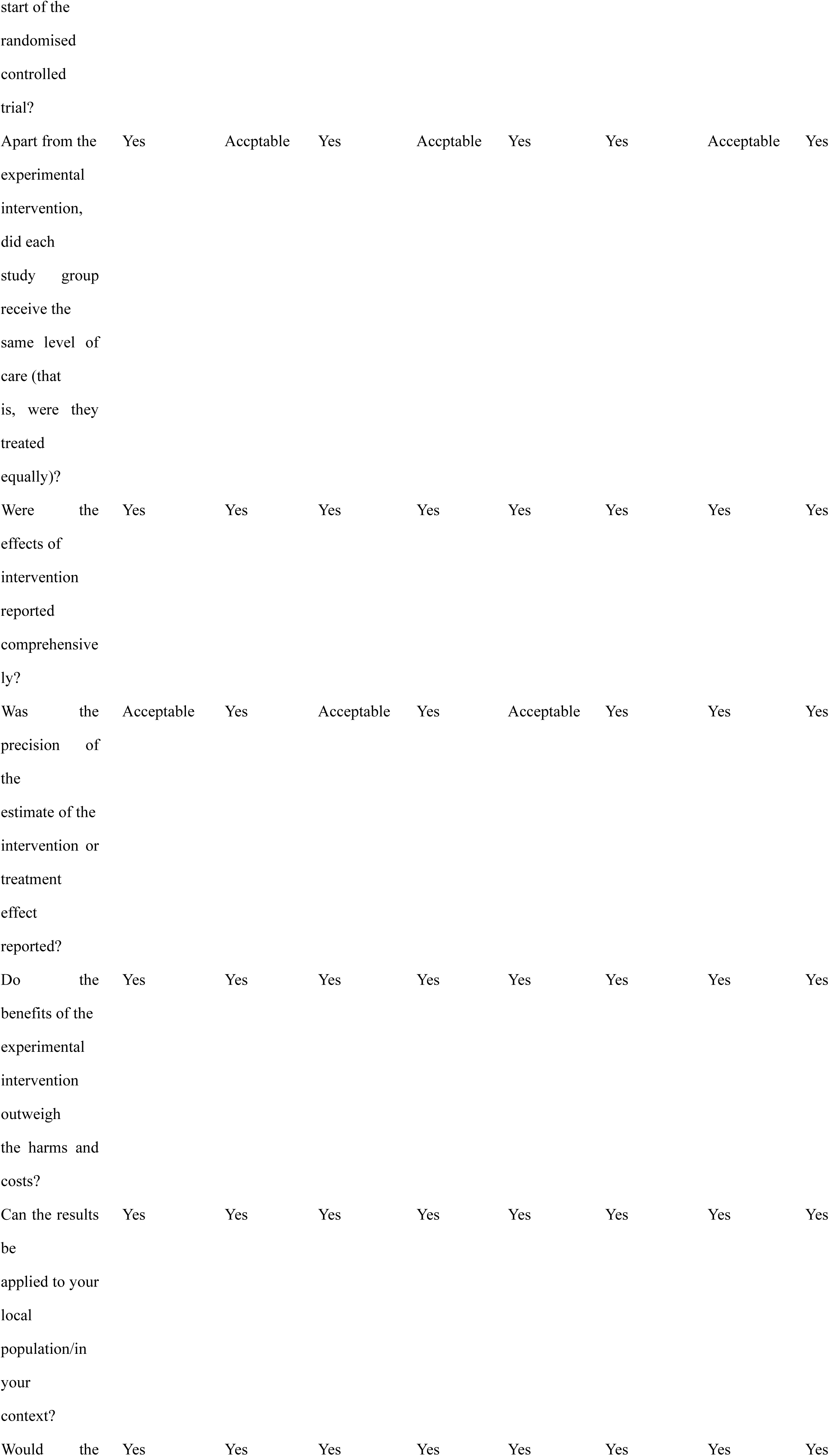
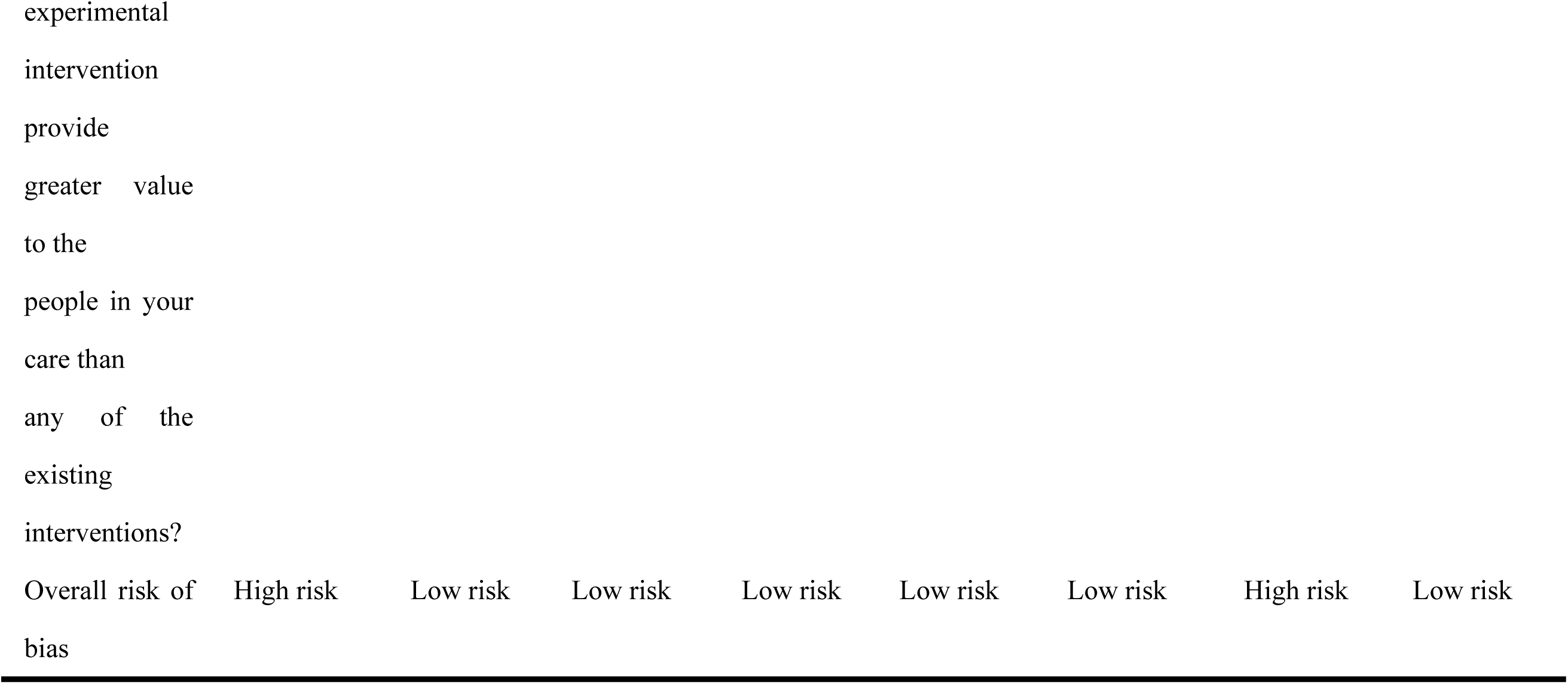
Critical Appraisal Skills Programme (CASP) quality assessment.

